# Iron Deficiency Anaemia in Mothers and Infants from a South African Birth Cohort: Prevalence and Profile in the Context of Inflammation

**DOI:** 10.1101/2025.01.23.25321033

**Authors:** Jessica E. Ringshaw, Michal R. Zieff, Sadeeka Williams, Chloë A. Jacobs, Zayaan Goolam Nabi, Thandeka Mazubane, Marlie Miles, Donna Herr, Khula South African Data Collection Team, Daniel C. Alexander, Melissa Gladstone, Vanja Klepac-Ceraj, Laurel J. Gabard-Durnam, Dima Amso, William P. Fifer, Derek K. Jones, Dan J. Stein, Steven C.R. Williams, Kirsten A. Donald

**Affiliations:** Department of Paediatrics and Child Health, Red Cross War Memorial Children’s Hospital, University of Cape Town, South Africa; Neuroscience Institute, University of Cape Town, South Africa; Centre for Neuroimaging Sciences, Department of Neuroimaging, Kings College London, United Kingdom; Centre for Medical Image Computing, Department of Computer Science, University College London, United Kingdom; Department of Women and Children’s Health, Institute of Life Course and Medical Science, Alder Hey Children’s NHS Foundation Trust, University of Liverpool, Liverpool, England, United Kingdom; Department of Biological Sciences, Wellesley College, Wellesley, Massachusetts, United States of America; Department of Psychology, Northeastern University, Boston, Massachusetts, United States of America; Department of Psychology, Columbia University, New York, New York, USA; Department of Psychiatry, Irving Medical Center, Columbia University, New York, New York, USA; Division of Developmental Neuroscience, New York State Psychiatric Institute, New York, New York, USA; Department of Pediatrics, Irving Medical Center, Columbia University, New York, New York, USA; Cardiff University Brain Research Imaging Centre, Cardiff University, Cardiff, Wales, United Kingdom; SA MRC Unit on Risk and Resilience in Mental Disorders, Department of Psychiatry & Neuroscience Institute, University of Cape Town, South Africa

**Keywords:** anaemia, iron deficiency, iron deficiency anaemia, inflammation, antenatal maternal health, child health, haemoglobin, mean corpuscular volume, serum ferritin, soluble transferrin receptor, highly sensitive C-Reactive Protein, Alpha-1 Acid Glycoprotein

## Abstract

**Objectives:** The scarcity of epidemiological data on anaemia in low- and middle-income countries, coupled with poor characterisation of overlapping risk factors in high-risk settings and contrasting approaches to the assessment of iron status with inflammation, represent critical gaps to address. This study aimed to characterise the prevalence and profile of iron deficiency anaemia, including adjustment for inflammation, in pregnant and postpartum women, as well as infants from South Africa.

**Methods:** Mother-child dyads (*n*=394) were recruited (2021-2022) for the Khula birth cohort study in Cape Town, South Africa. Haematological metrics (haemoglobin, mean corpuscular volume [MCV]), iron metrics (serum ferritin and soluble transferrin receptor [sTfR]), and inflammatory biomarkers (highly sensitive C-Reactive Protein [*hs*CRP]; Alpha-1 Acid Glycoprotein [AGP]) were obtained from mothers antenatally and postnatally, as well as from infants 3-18 months after birth. World Health Organisation (WHO) guidelines were used to classify anaemia and iron deficiency. The extent to which inflammation impacted iron deficiency was assessed using two methods: Method A: higher serum ferritin thresholds for classifying iron status in participants with inflammation (WHO), Method B: Biomarkers Reflecting Inflammation and Nutritional Determinants of Anaemia (BRINDA) regression which corrects serum ferritin levels based on inflammatory biomarker concentrations.

**Results:** Prevalence of anaemia was 34.74% (107/308) in pregnancy and 22.50% (54/240) in mothers at 3-6 months postpartum. Of their infants, 46.82% (125/267) and 48.10% (136/283) were anaemic at least once by 6-12 months and 12-18 months, respectively. When accounting for inflammation using Method A, the prevalence of maternal iron deficiency (regardless of anaemia), increased from 18.35% (20/109) to 55.04% (60/109) in pregnancy, and from 11.97% (28/234) to 46.58% (109/234) postnatally. Similarly, using Method B, the estimated prevalence of maternal iron deficiency increased to 38.53% (42/109) in pregnancy, and 25.21% (59/234) postnatally. In infants at 12-18 months, the prevalence of iron deficiency increased from 19.79% (19/96) to 31.25% (30/96) and 32.29% (31/96) using Methods A and B, respectively. Approximately half of anaemia cases in mothers antenatally (50%; 20/40) and postnatally (45.10%; 23/51), and infants at 12-18 months (55.56%; 10/18), were attributable to iron deficiency. However, there was little overlap in the estimated prevalence of microcytic anaemia (based on MCV) and iron deficiency anaemia (based on adjusted serum ferritin) in pregnant and postpartum mothers, as well as infants at 3-6 and 6-12 months. At these timepoints, microcytic anaemia underestimated the likely proportion of anaemia cases attributable to iron deficiency.

**Conclusion:** This is one of the first studies to report the true prevalence of iron deficiency anaemia in South African mothers and infants, and the extent to which it may be underestimated if inflammation is not accounted for. Additionally, the results indicate that, while microcytic anaemia classification may be a valid proxy for iron deficiency anaemia in infants over 1 year of age, it seems less useful for pregnant and postpartum mothers and younger infants within the context of inflammation. Overall, the findings contribute to a global effort to understand the complex aetiology of iron deficiency anaemia, informing guidelines for optimised detection, prevention, and intervention in high-risk communities.

## INTRODUCTION

Anaemia, a haematological disorder diagnosed by low serum haemoglobin, is estimated to affect 1.8 billion people worldwide, with women and children being the most susceptible [1–3]. This public health concern is prominent in low- and middle-income countries (LMICs) where the prevalence of anaemia is disproportionately high due to various contributing factors such as poverty, food insecurity, malnutrition, and infectious disease [4–7]. In comparison to high-income countries where 15% of pregnant women and 10% of young children (6-59 months) are estimated to be anaemic, corresponding statistics in Africa and South Asia range between 30-50% and 35-55% respectively [1].

Concurrent with an increased focus on antenatal maternal anaemia as an important driver of poor developmental outcomes [8], novel neuroimaging research has provided evidence for an association between maternal anaemia in pregnancy and structural child brain alterations [9] that persist with age [10]. This supports a growing need to understand the aetiology of this complex condition, and to provide deeper insight into the profile and burden of anaemia in LMICs. In recognising anaemia as a priority for targeted prevention and intervention, it has been included in policy briefs [11, 12] including the World Health Organisation (WHO) Global Nutrition Targets 2025 [12]. However, efforts to reduce the prevalence of anaemia stagnated between 2000 and 2019 [1], with progress being made at half the pace of other nutritional indicators for maternal and child health [13]. Consequently, and in alignment with the United Nations (UN) Sustainable Development Goals (SDGs) [14], the WHO and UN International Emergency Fund (UNICEF) proposed an extension of nutrition targets to 2030 [15] and developed a framework for accelerated anaemia reduction [2].

This WHO framework emphasizes the complex aetiology of anaemia, identifying the main direct causes as micronutrient deficiencies, chronic infection and inflammation, gynaecological and obstetric conditions, and inherited blood disorders [2]. Given that iron deficiency is reported as accounting for more than 50% of anaemia cases, it has been the most common focus in previous work. Iron deficiency anaemia (IDA) occurs when haemoglobin production is limited by a chronic negative iron balance, representing either an absolute or functional iron deficiency [16]. Absolute iron deficiency is characterized by a total reduction in iron stores due to increased demand, decreased intake, malabsorption, or chronic blood loss [17–19]. While decreased intake is typically due to poor nutrition, instances of increased demand include pregnancy, and malabsorption may be attributable to other factors including alcohol use. In contrast, functional iron deficiency refers to adequate iron stores but insufficient bioavailability of iron. This may be observed in disease pathologies associated with inflammation due to increased iron sequestration [17–19]. Therefore, in communities with an overlapping high prevalence of nutritional risk, substance use, and infectious disease such as HIV, the profile of IDA may be complex and require more nuanced assessment and intervention strategies.

In order to effectively reduce the prevalence of anaemia, the WHO recommends five action areas with the first being to analyse data on direct causes and underlying risk factors for anaemia [2]. In addition to assessing anaemia using haemoglobin measures, other data including iron metrics and inflammatory biomarkers are important in establishing the context-specific drivers and local risk factors for anaemia. Metrics typically used to assess iron status, such as serum ferritin and soluble transferrin receptor (sTfR), are altered by inflammatory mechanisms associated with the body’s acute-phase response to infection [20–22]. To account for this, approaches include adjusted serum ferritin thresholds for confirming iron deficiency where there is evidence of inflammation (WHO) [23] or the use of regression models to correct continuous concentrations for iron status metrics (serum ferritin and sTfR) based on inflammatory biomarker concentrations [24, 25]. The latter has been developed by Biomarkers Reflecting Inflammation and Nutritional Determinants of Anemia (BRINDA) [26], an ongoing international collaboration to inform global anaemia guidelines on micronutrient assessment and anaemia characterisation [27]. While efforts to refine the BRINDA model are ongoing, reproducible work using this correction approach has demonstrated higher estimates of iron deficiency in children and non-pregnant women after accounting for inflammation [24]. It is particularly relevant in countries with a high burden of infection [28], with findings separately demonstrated across Africa in children with Malaria [29, 30] and pregnant women [31].

However, there is an ongoing need for further research on a broader range of infectious and pro-inflammatory exposures. This may be particularly useful in South Africa where 23.5% of women (aged 15-49) are estimated to be living with HIV, representing one of the highest prevalences worldwide [32]. Furthermore, global population research estimating the prevalence of iron deficiency in children and women of reproductive age with adjustment for inflammation do not include pregnancy data [33]. Therefore, very little is known regarding the typical reference ranges for iron metrics across trimesters. Notably, South Africa is not represented in this pooled population analysis, highlighting an even greater need for contributions from this context. This is particularly relevant given that most references ranges for anaemia and iron-deficiency are derived from iron-supplemented women in high-income countries, and may be inappropriate for use in communities of African descent with a genetic predispositions for naturally lower concentrations [3].

Given the relative lack of epidemiological data in LMICs [1, 33], the poor characterisation of context-specific overlapping risk factors including iron deficiency in high-risk communities [2], and limitations in the assessment of biomarkers for iron status within the context of inflammation [24], precise reporting using comprehensive data on iron deficiency and anaemia is needed. Furthermore, the widespread use of microcytic anaemia classification (based on mean corpuscular volume; MCV) as a proxy for iron deficiency anaemia in clinical practice [34] requires further validation, particularly in settings with a high burden of infection, through comparison with iron metrics that have been adjusted for inflammation.

The aim of this birth cohort study is to contribute towards a global effort to characterise the prevalence and profile of IDA in pregnant women, postpartum women, and infants using haematological metrics, iron metrics, and inflammatory biomarkers from South Africa.

## METHODS

### Study Design and Setting

This report is based on the Khula South Africa Study, an observational population-based birth cohort [35]. Pregnant women were recruited antenatally from Gugulethu Midwife Obstetrics Unit and postpartum from nearby clinics in the area. Gugulethu is an urban informal settlement at sea level in the Western Cape Province and is populated by a community at high risk of malnutrition and HIV. All mother-child dyads were followed prospectively with postnatal study visits conducted at the University of Cape Town Neuroscience Institute.

### Participants

South African women over the age of 18 years were recruited (6^th^ December 2021 – 29^th^ November 2022) during their third trimester of pregnancy (28-40 weeks) or up to 3-6 months postpartum. Exclusion criteria included multiple pregnancies, psychotropic drug use during pregnancy, infant congenital malformations or abnormalities (e.g. Spina Bifida, Down’s Syndrome), and severe birth complications (e.g. uterine rupture, birth asphyxia). Overall, a total of 394 mother-child pairs from South Africa were enrolled in the Khula Study, with 329 women recruited antenatally and 65 women recruited postpartum (Figure 1). Antenatal and postnatal (first study visit; approximately 3-6 months postpartum) maternal anaemia and iron metrics data was acquired for a sub-group of mothers from study samples or extracted from the National Health Laboratory Services (NHLS) database (records accessed: 8^th^ April 2024 – 12^th^ April 2024). Child anaemia and iron metrics data were collected across 3 study visits postnatally (at approximately 3-6 months, 6-12 months, and 12-18 months) in early life.

**Figure 1.**
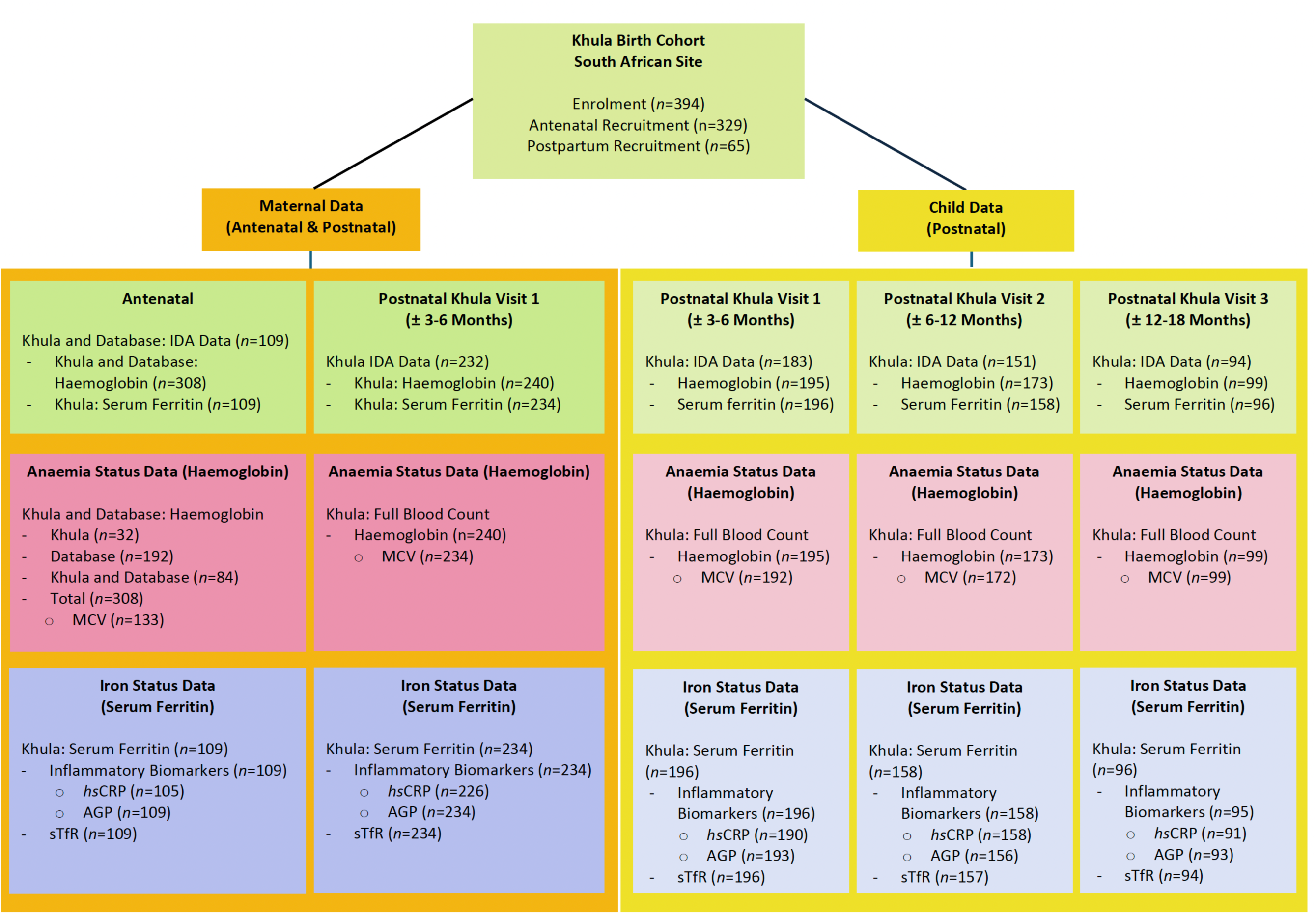
Khula Study Flow Chart Demonstrating Maternal and Child Data Available Across Timepoints for the Assessment of Iron Deficiency Anaemia (IDA) in South Africa Abbreviations: MCV = Mean Corpuscular Volume, *hs*CRP = Highly Sensitive C-Reactive Protein, AGP = Alpha-1Acid Glycoprotein, sTfR = Soluble Transferrin Receptor

Maternal and infant data across measures and timepoints is represented in the study flowchart (Figure 1). All mothers provided written informed consent at enrolment and were re-consented annually. While specified, approved authors had access to identifiable participant information during data collection, only non-identifiable data was captured and analyzed.

### Measures

#### Contextual Measures

Demographic, psychosocial, and medical information was collected antenatally and postnatally for contextual characterisation of the sample. Self-reported maternal HIV status during pregnancy and child HIV status were confirmed by data from the Provincial Health Data Centre (PHDC), with mothers tested during pregnancy as per national policy. Antenatal maternal alcohol exposure was dichotomously classified at enrolment based on reported consumption of more than two drinks twice per week in any trimester (Alcohol Exposure Questionnaire; AEQ), or high risk of dependence in the preceding 3 months (score >12) using the Alcohol, Smoking, and Substance Involvement Screening Test (ASSIST) [36, 37].

Tobacco use was also coded dichotomously based on any use reported over the previous three months using the ASSIST. For antenatally-enrolled mothers, this three-month period corresponded to the second or third trimester of pregnancy. Maternal depression was dichotomised based on moderate-severe symptomology (score >12) using the Edinburgh Postnatal Depression Scale (EPDS) [38] at enrolment. A food insecurity proxy was obtained at enrolment using items assessing food resource sufficiency in the last year (question 38 and 39) from the Life Events Questionnaire, a tool adapted from the WHO World Mental Health Survey [39]. The exacerbating effect of the COVID-19 pandemic on food insecurity was assessed using an adapted version of the COPE-IS: Coronavirus Perinatal Experiences –

Impact Survey [40]. Child anthropometry was obtained by trained medical doctors on the research team at neuroimaging study visits. Child head circumference, weight, and length/height measurements were converted to z-scores based on age and sex using Anthro software for WAZ, HAZ, and HCZ. Infants were classified as underweight, stunted, or microcephalic if they had z-scores of less than −2 standard deviations.

#### Anaemia

Antenatal maternal anaemia was assessed based on serum haemoglobin from a full blood count (FBC) in pregnancy (at Khula enrolment), or extracted from the NHLS database. For mothers with multiple measurements from either source, the lowest haemoglobin value and corresponding gestational age at the time of measurement was selected to represent the timepoint for most severe anaemia. Antenatal maternal anaemia status was dichotomously classified using recently updated (2024) WHO thresholds of <11g/dL in trimesters 1 and 3, and <10.5g/dL in trimester 2 [41]. Further severity categorisations were made indicating mild (haemoglobin 10.0 – 10.9g/dL in trimester 1 and 3; 9.5 – 10.4g/dL in trimester 2), moderate (haemoglobin 7.0 – 9.9g/dL in trimester 1; 7 – 9.4 g/dL in trimester 2), and severe (haemoglobin <7.0g/dL) anaemia. For mothers for whom gestational age was not indicated at the time of measurement, haemoglobin was assigned to be from trimester 3 for classification given that this was when mothers were recruited. This is also when the physiological risk of anaemia is greatest as iron demand initially drops in pregnancy due to the cessation of menstruation, but rapidly increases with foetal growth [3, 16]. Postnatal maternal anaemia was based on serum haemoglobin from an FBC at the first study visit (approximately 3-6 months postpartum) using the WHO threshold for non-pregnant women (haemoglobin <12g/dL) [41]. Severity was classified as mild (11 – 11.9g/dL), moderate (8 – 10.9g/dL), and severe (<8g/dL). Haemoglobin measurements were not corrected for smoking or altitude given the extremely low prevalence of maternal tobacco use and the fact that the community lives at sea level. Self-reported maternal anaemia status was recorded at enrolment for comparison with observed prevalence.

Child anaemia at study visit 1, 2, and 3 was dichotomously classified according to age-specific cut-offs using WHO guidelines [42] for children over 6 months and local guidelines for children under 6 months (Groote Schuur Hospital/University of Cape Town Pathology Laboratory; Supplementary Information, Table S1). Classifications across timepoints were used to determine the percentage of infants diagnosed with anaemia at least once by study visit 2 (within the first 6-12 months) and study visit 3 (within the first 12-18 months).

Mean corpuscular volume (MCV), a measure of average red blood cell size and volume, was acquired from FBCs for mothers and infants. Given the widespread use of MCV in clinical practice as a proxy for anaemia aetiology, values were categorised accordingly with microcytic anaemia (below average MCV) considered likely to represent iron-deficiency anaemia, macrocytic anaemia (larger than average MCV) often being linked to pernicious anaemia (lack of limited absorption of vitamin B12) or folic acid deficiency, and normocytic anaemia (normal MCV) mostly considered to be associated with chronic infection, blood loss or kidney failure [34]. Antenatal maternal MCV measures were classified as microcytic using trimester-specific thresholds (<81µm^3^ in trimester 1, <82 µm^3^ in trimester 2; <81 µm^3^ in trimester 3) [43]. For mothers with no gestational age at the time of measurement, MCV was assigned to trimester 3. Postnatal maternal MCV was classified as microcytic if <79µm^3^ [34]. Based on Centers for Disease Control (CDC) paediatric guidelines for the first year of life, infant MCV values were classified as microcytic if <77µm^3^ [44].

#### Inflammation

Inflammation in mothers and infants was assessed using two well-known acute-phase response proteins, namely highly sensitive C-Reactive Protein (*hs*CRP) and Alpha-1Acid Glycoprotein (AGP) [45]. These were used as biomarkers of inflammation (dichotomous classification) with *hs*CRP >5mg/L and AGP >1g/L being indicative of acute and chronic inflammation, respectively. These thresholds are consistent with WHO guidelines on interpreting serum ferritin [23, 46] and BRINDA methodology for adjusting iron status [24].

#### Iron Deficiency

Iron deficiency was assessed using iron metrics including serum ferritin and sTfR, with adjustment based on the inflammatory biomarkers. Serum ferritin, a protein reflecting body iron stores, was used to classify iron deficiency status before adjusting for inflammation based on WHO thresholds of <12μg/L in children under five years of age, and <15μg/L in women of reproductive age (including during pregnancy due to lack of defined thresholds for this period) [23]. However, given that serum ferritin is an acute-phase reactant known to be transiently elevated in states of inflammation, two adjustment methods were applied to obtain and compare corrected classification of iron status. For Method A, higher adjusted WHO thresholds were used for participants with positive biomarkers of inflammation (*hs*CRP >5mg/L or AGP >1g/L) [23, 46]. In these instances, serum ferritin concentrations of <30μg/L and <70μg/L were used to classify iron deficiency for children under five years of age and women of reproductive age, respectively. Method B was based on the BRINDA regression modelling methodology [24] using a statistical package in R (BRINDA: Computation of BRINDA Adjusted Micronutrient Biomarkers for Inflammation; R package version 0.1.5) [47] to correct continuous serum ferritin concentrations based on *hs*CRP and AGP concentrations. Standard WHO thresholds of <12μg/L and <15μg/L were subsequently used to classify iron deficiency based on the adjusted serum ferritin concentrations in infants and women of reproductive age, respectively [23].

In addition to serum ferritin, sTfR was also used as a complementary iron metric as it represents the body’s demand for iron, with the over-expression of this blood protein suggesting iron-deficient erythropoiesis. While sTfR is not an acute-phase reactant and is considered to be a stable measure for iron status in instances of inflammation [48, 49], there is some evidence to suggest the need for adjustment based on AGP [25]. It is proposed that inflammation may contribute to an increase in the transferrin receptor expression due to the redistribution of iron [22], resulting in the overestimation of iron-deficient erythropoiesis when AGP is high [25]. Therefore, the BRINDA regression correction approach was used to adjust sTfR concentrations in R [47]. Given that there are no standardised universal reference ranges for sTfR [46], a cutoff of >8.3mg/L was used to support a diagnosis of iron deficiency for women and infants. This was based on laboratory-specific guidelines (VitMinLab) [50] for the sandwich ELIZA technique [51] which is calibrated to commercial Ramco assay kits. This was applied to unadjusted and adjusted sTfR concentrations, and is consistent with previous BRINDA methodology in research on women and children [24].

### Statistical Analysis

Sample characteristics (demographic, maternal exposures, anthropometric, and food insecurity) were presented as means and standard deviations for continuous variables and frequencies for categorical variables. All statistical analyses were conducted in SPSS (Version 29). A two-sided significance level of *p*<0.05 was used throughout.

For the first dimension of results (haemoglobin metrics), the self-reported prevalence of maternal anaemia at enrolment was recorded and the actual prevalence and severity of maternal anaemia were determined for the antenatal and postnatal timepoints. Similarly, child anaemia prevalence was documented at each study visit (1-3) separately, with reporting of incidence within the first 6-12 months (by study visit 2) and 12-18 months of life (by study visit 3). To characterize the haemoglobin profile of women and infants in this population sample, the mean, standard deviation, and range of values were reported for anaemic and non-anaemic participants. Group differences in sociodemographic and clinical characteristics based on antenatal maternal anaemia and child anaemia classification within the first 12-18 months of life were determined using independent samples t-tests for continuous data and chi-squared or Fisher’s exact test for categorical data. To assess whether antenatal maternal anaemia increases the risk of postnatal maternal anaemia or child anaemia, the association between antenatal and postnatal anaemia status for mothers and infants was calculated using Pearson correlation coefficient.

For the second dimension of results (iron metrics), the prevalence of inflammation was determined for mothers and infants, and associations between inflammatory and iron biomarkers was assessed using Pearson correlation coefficient. For both Methods A and B (higher WHO thresholds versus BRINDA corrected serum ferritin concentrations), unadjusted and adjusted estimates of iron deficiency were represented across timepoints, with the percentage point change reported after accounting for inflammation. Given that the BRINDA approach adjusts serum ferritin concentrations on an individual level using corresponding inflammatory markers, it is considered to be more specific than general threshold adjustments. Therefore, sample characteristics across groups were reported based on antenatal maternal iron deficiency classification with adjustment for inflammation using the BRINDA approach.

For the third dimension of results (IDA metrics), the overall summary statistics for mothers and infants with IDA (low haemoglobin and low serum ferritin) using adjusted metrics were reported. Similarly, for all anaemic mothers and infants with MCV and adjusted serum ferritin data, further reporting was conducted to determine the prevalence of microcytic anaemia in this sample, and its relative overlap with IDA.

## RESULTS

The results are described across three dimensions, namely haemoglobin metrics, iron metrics, and IDA metrics. Firstly, haemoglobin metrics were assessed to report the prevalence of anaemia and to characterise a typical haematological profile in this sample. Secondly, iron metrics were described in reporting the prevalence of iron deficiency in the cohort, both with and without adjustment for inflammation. Thirdly, in anaemic mothers and infants, iron and MCV metrics were used to report on the comparative prevalence and overlap between IDA and microcytic anaemia classification.

### Dimension 1: Haemoglobin Metrics

The first dimension of the results describes the profile and severity of anaemia for mothers and infants with low haemoglobin, based on WHO thresholds for classification in this cohort. It also summarises haemoglobin metrics in a population sample (i.e. South African mothers and infants without anaemia) as a standard for reference (mean, standard deviation, range).

This is described for mothers in pregnancy and postpartum, as well as for infants postnatally at study visits 1 (approximately 3-6 months), 2 (approximately 6-12 months), and 3 (approximately 12-18 months). Sample characteristics across groups (anaemic versus non-anaemic) are detailed to explore potential differences that may be contributing to the risk profile for anaemia in this context.

#### Antenatal Maternal Anaemia

In this sample of mothers (Table 1; Mean [SD] age of 28.84 [5.66] years; age range of 18 – 41.70 years), haemoglobin was available either from Khula FBCs at enrolment, or the NHLS database. Minimum haemoglobin concentrations were predominantly measured in the second (102/241[42.32%]) and third (86/241[35.68%]) trimester, at a median (IQR) gestation of 22 (14.00 - 31.50) weeks. In this sample, 34.74% (107/308) of mothers were anaemic in pregnancy, of which 63.55% (68/107) had mild anaemia, 34.58% (37/107) had moderate anaemia, and 1.87% (2/107) had severe anaemia. However, only 4/107 (3.74%) anaemic mothers correctly self-reported a positive diagnosis of anaemia during pregnancy (Table 1).

**Table 1.** Maternal and Infant Sample Characteristics According to Antenatal Maternal Anaemia Status.

| Variable<sup>a</sup> | Total Sample (*n* = 308) |  | *p* |
| --- | --- | --- | --- |
|  | Maternal Anaemia<br>(*n* = 107) | No Maternal Anaemia<br>(*n* = 201) |  |
| **Antenatal Maternal Anaemia Profile** |  |  |  |
| Self-reported anaemia in pregnancy <sup>f,g</sup> | 4 (3.74) | 1 (0.50) | 0.037* |
| Actual anaemia status in pregnancy <sup>b</sup> |  |  |  |
| Mild | 68 (63.55) | n/a |  |
| Moderate | 37 (34.58) | n/a |  |
| Severe | 2 (1.87) | n/a |  |
| Gestational age at Hb measurement <sup>g</sup> | 26.70 (9.21)<br>[4 – 40] | 20.72 (9.81)<br>[5 - 41] | <0.001*** |
| Pregnancy trimester Hb measured <sup>c,g</sup> |  |  |  |
| First | 11 (10.28) | 42 (20.90) |  |
| Second | 26 (24.30) | 76 (37.81) | <0.001*** |
| Third | 42 (39.25) | 44 (21.89) |  |
| Minimum Hb in pregnancy (g/dL) <sup>c</sup> | 9.76 (1.07)<br>[4.4 – 10.9] | 11.85 (0.74)<br>[10.5 – 14.0] | <0.001*** |
| Trimester 1 (*n*=53) | 10.18 (0.99)<br>[7.9 – 10.9] | 12.16 (0.70)<br>[11.0 – 13.5] | <0.001*** |
| Trimester 2 (*n*=102) | 9.72 (0.61)<br>[7.7 – 10.3] | 11.73 (0.77)<br>[10.5 – 13.8] | <0.001*** |
| Trimester 3 (*n*=86) <sup>e</sup> | 9.64 (1.35)<br>[4.4 – 10.9] | 11.67 (0.59)<br>[11.0 – 13.5] | <0.001*** |
| **Maternal Characteristics** |  |  |  |
| Monthly household income (ZAR) <sup>f,g</sup> |  |  |  |
| <1000 | 20 (18.69) | 34 (16.92) |  |
| 1000-5000 | 57 (53.27) | 82 (40.80) | 0.060 |
| 5000-10000 | 22 (20.56) | 51 (25.37) |  |
| >10000 | 1 (0.93) | 12 (5.97) |  |
| Education <sup>f</sup> |  |  |  |
| Primary | 4 (3.74) | 2 (1) |  |
| Some secondary | 84 (78.50) | 55 (27.36) |  |
| Completed secondary | 84 (78.50) | 37 (18.41) | 0.609 |
| Some tertiary | 16 (14.95) | 8 (3.98) |  |
| Completed tertiary | 13 (12.15) | 5 (2.49) |  |
| Employed <sup>g</sup> | 34 (31.78) | 74 (36.82) | 0.361 |
| Age at enrolment (years) <sup>g</sup> | 28.93 (5.65)<br>[18.5 - 40.5] | 28.79 (5.69)<br>[18.0 – 41.7] | 0.843 |
| Food insecurity | 55 (51.40) | 119 (59.20) | 0.188 |
| COVID-19 effect on food insecurity <sup>g</sup> |  |  |  |
| No disruption | 37 (34.58) | 85 (42.29) |  |
| Some disruption | 12 (11.21) | 34 (16.92) | 0.470 |
| Extreme disruption | 17 (15.89) | 28 (13.93) |  |
| Smoking during pregnancy <sup>f</sup> | 7 (6.54) | 5 (2.49) | 0.119 |
| Alcohol during pregnancy | 10 (9.35) | 9 (4.48) | 0.091 |
| Depression during pregnancy | 18 (16.82) | 34 (16.92) | 0.983 |
| HIV infection during pregnancy <sup>g</sup> | 47 (43.93) | 57 (28.36) | 0.005** |
| **Infant Characteristics at Birth** |  |  |  |
| Sex (male) <sup>g</sup> | 37 (34.58) | 93 (46.27) | 0.249 |
| Gestational age at birth (weeks) <sup>g</sup> | 38.85 (1.87) | 39.22 (1.86) | 0.145 |
|  | [32 - 42] | [30 - 42] |  |
| HIV infection<sup>f,g</sup> | 1 (0.93) | 0 (0) | 0.439 |
| Birth weight (g)<sup>d,g</sup> | 3104.93 (484.49) | 3222.01 (504.89) | 0.077 |
|  | [1950 - 4360] | [1180 - 4750] |  |
| Birth length (cm)<sup>d,g</sup> | 49.33 (3.46) | 50.14 (3.40) | 0.075 |
|  | [41 - 57] | [33 - 57] |  |
| Birth head circumference (cm)<sup>d,g</sup> | 34.58 (1.58) | 34.65 (1.79) | 0.783 |
|  | [30.5 - 38] | [28 - 40] |  |

Abbreviations: Hb, haemoglobin; ZAR, South African Rand; HIV, Human Immunodeficiency Virus; COVID-19, Coronavirus Disease 2019; g, grams; cm, centimetres.
SI conversion factor: To convert to haemoglobin grams per litre, multiply by 10.
<sup>a</sup> Values for continuous variables are presented as: mean (standard deviation) [range]. Values for categorical variables are presented as: count (%).
<sup>b</sup> Maternal anaemia during pregnancy was classified according to the WHO threshold of <11g/dL for trimesters 1 and 3, and <10.5g/dL for trimester 2. Severity categorisations were made indicating mild (haemoglobin 10.0 – 10.9g/dL in trimester 1 and 3; 9.5-10.4 in trimester 2), moderate (haemoglobin 7.0 – 9.9g/dL in trimester 1; 7-9.4 in trimester 2), and severe (haemoglobin <7.0g/dL) anaemia categories.
<sup>c</sup> Trimester of pregnancy defined as first (0-12 weeks), second (13-27 weeks), and third (28 weeks onwards).
<sup>d</sup> The birth anthropometric measurements were conducted by trained labour staff in the ward. Infant length was measured in cm to the nearest completed 0.5cm and weight was measured in kgs.
<sup>e</sup> Levene's test was significant. *T*-test results were interpreted based on equal variance not assumed.
<sup>f</sup> Fisher's exact test result interpreted due to one or more cells having an expected count of less than 5.
<sup>g</sup> Missing values: self-reported anaemia in pregnancy (*n* = 44), gestational age at maternal minimum haemoglobin measurement (*n* = 67), pregnancy trimester Hb measured (*n* = 67), monthly household income (*n* = 29), maternal employment (*n* = 1), maternal age at enrolment (*n* = 67), COVID-19 effect on food insecurity (*n* = 95), maternal HIV infection during pregnancy (*n* = 5), infant sex (*n* = 44), infant gestational age at birth (*n* = 67), infant HIV infection (*n* = 226), infant birth weight (*n* = 46), infant birth length (*n* = 50), infant birth head circumference (*n* = 49).
\**p* is significant at <0.05, \*\**p* is significant at <0.01, \*\*\**p* is significant at <0.001.

There were no group differences in maternal and infant sample characteristics between anaemic and non-anaemic mothers, with the exception of maternal HIV infection which was significantly more prevalent in mothers with anaemia (47/107 [43.93%]) than mothers without anaemia (57/201 [28.36%]) in pregnancy. While smoking and alcohol use in pregnancy were not common, the prevalence of reported food insecurity was high in both mothers with antenatal anaemia (55/107 [51.40%]) and mothers without (119/201 [59.20%]). Additionally, the effect of the COVID-19 pandemic on food insecurity was similar with approximately one third of both groups reporting some or extreme disruption.

#### Postnatal Maternal Anaemia

At study visit 1 (approximately 3-6 months postpartum), 240 mothers (mean [SD] age of 28.95 [5.59] years; range of 18-44 years) had haemoglobin measures from Khula FBCs (Table 2). Overall, 22.5% (54/240) of mothers were found to be anaemic postpartum, of which 75.93% (41/54) had mild anaemia, 20.37% (11/54) had moderate anaemia, and 3.70% (2/54) had severe anaemia. As seen in the group of mothers with antenatal maternal haemoglobin data, there were no group differences in maternal or infant sample characteristics at study visit 1, with the exception of maternal HIV infection which was more prevalent in mothers with postnatal anaemia (50% [27/54]) than mothers without postnatal anaemia (31.18% [58/186]; Table 2).

**Table 2.**
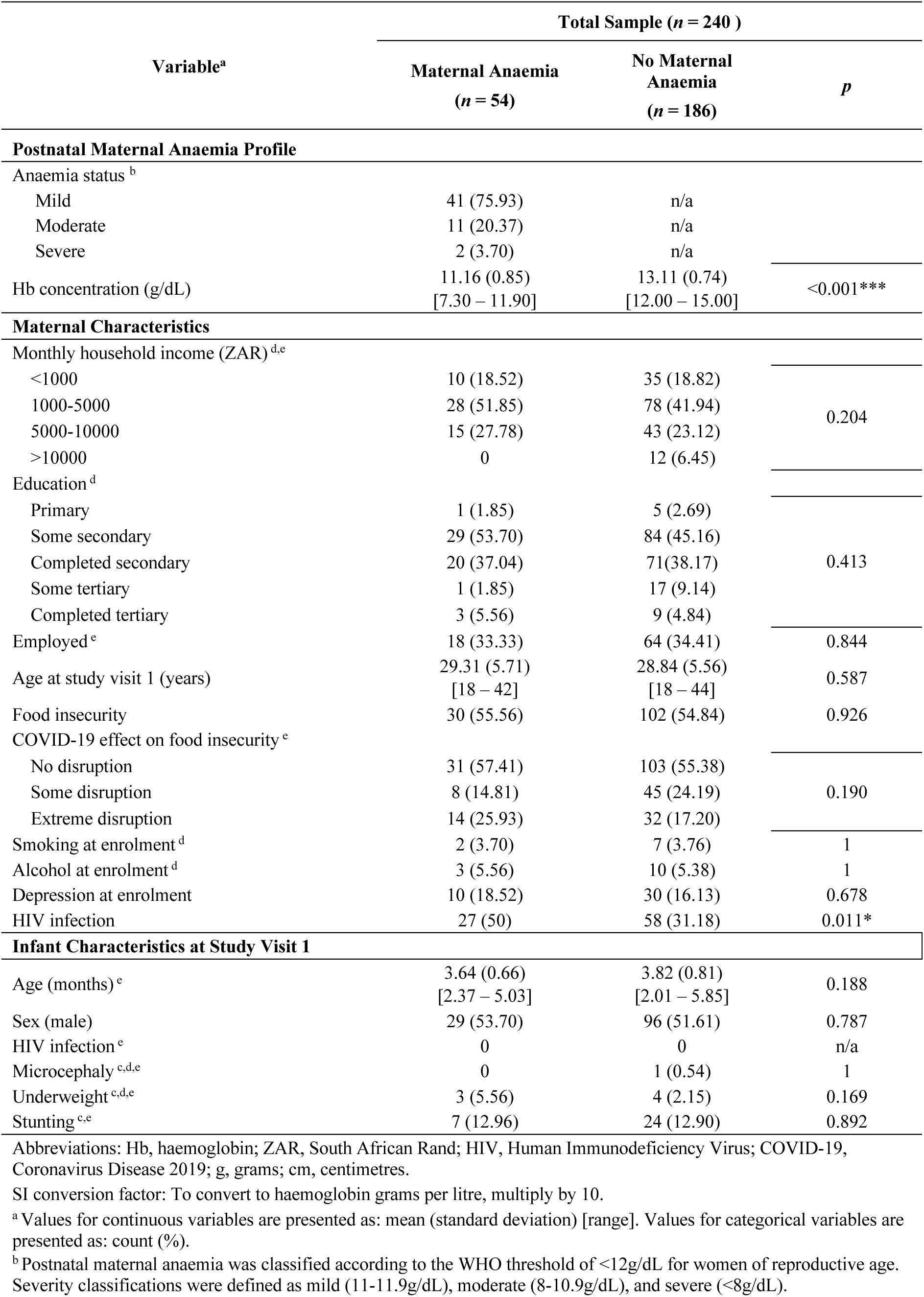
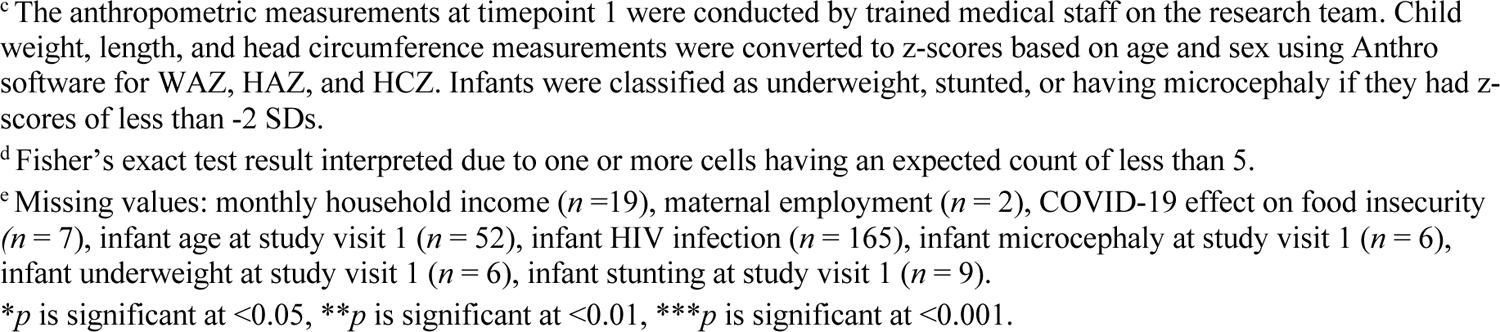
Maternal and Infant Sample Characteristics According to Postnatal Maternal Anaemia Status at Study Visit 1 (±3-6 Months Postpartum)

| Variable<sup>a</sup> | Total Sample (n = 240 ) |  |  |
| --- | --- | --- | --- |
|  | Maternal Anaemia<br>(n = 54) | No Maternal<br>Anaemia<br>(n = 186) | *p* |
| Postnatal Maternal Anaemia Profile |  |  |  |
| Anaemia status <sup>b</sup> |  |  |  |
| Mild | 41 (75.93) | n/a |  |
| Moderate | 11 (20.37) | n/a |  |
| Severe | 2 (3.70) | n/a |  |
| Hb concentration (g/dL) | 11.16 (0.85)<br>[7.30 – 11.90] | 13.11 (0.74)<br>[12.00 – 15.00] | <0.001*** |
| Maternal Characteristics |  |  |  |
| Monthly household income (ZAR) <sup>d,e</sup> |  |  |  |
| <1000 | 10 (18.52) | 35 (18.82) | 0.204 |
| 1000-5000 | 28 (51.85) | 78 (41.94) |  |
| 5000-10000 | 15 (27.78) | 43 (23.12) |  |
| >10000 | 0 | 12 (6.45) |  |
| Education <sup>d</sup> |  |  |  |
| Primary | 1 (1.85) | 5 (2.69) | 0.413 |
| Some secondary | 29 (53.70) | 84 (45.16) |  |
| Completed secondary | 20 (37.04) | 71(38.17) |  |
| Some tertiary | 1 (1.85) | 17 (9.14) |  |
| Completed tertiary | 3 (5.56) | 9 (4.84) |  |
| Employed <sup>e</sup> | 18 (33.33) | 64 (34.41) | 0.844 |
| Age at study visit 1 (years) | 29.31 (5.71)<br>[18 – 42] | 28.84 (5.56)<br>[18 – 44] | 0.587 |
| Food insecurity | 30 (55.56) | 102 (54.84) | 0.926 |
| COVID-19 effect on food insecurity <sup>e</sup> |  |  |  |
| No disruption | 31 (57.41) | 103 (55.38) | 0.190 |
| Some disruption | 8 (14.81) | 45 (24.19) |  |
| Extreme disruption | 14 (25.93) | 32 (17.20) |  |
| Smoking at enrolment <sup>d</sup> | 2 (3.70) | 7 (3.76) | 1 |
| Alcohol at enrolment <sup>d</sup> | 3 (5.56) | 10 (5.38) | 1 |
| Depression at enrolment | 10 (18.52) | 30 (16.13) | 0.678 |
| HIV infection | 27 (50) | 58 (31.18) | 0.011* |
| Infant Characteristics at Study Visit 1 |  |  |  |
| Age (months) <sup>e</sup> | 3.64 (0.66)<br>[2.37 – 5.03] | 3.82 (0.81)<br>[2.01 – 5.85] | 0.188 |
| Sex (male) | 29 (53.70) | 96 (51.61) | 0.787 |
| HIV infection <sup>e</sup> | 0 | 0 | n/a |
| Microcephaly <sup>c,d,e</sup> | 0 | 1 (0.54) | 1 |
| Underweight <sup>c,d,e</sup> | 3 (5.56) | 4 (2.15) | 0.169 |
| Stunting <sup>c,e</sup> | 7 (12.96) | 24 (12.90) | 0.892 |

Abbreviations: Hb, haemoglobin; ZAR, South African Rand; HIV, Human Immunodeficiency Virus; COVID-19, Coronavirus Disease 2019; g, grams; cm, centimetres.
SI conversion factor: To convert to haemoglobin grams per litre, multiply by 10.
<sup>a</sup> Values for continuous variables are presented as: mean (standard deviation) [range]. Values for categorical variables are presented as: count (%).
<sup>b</sup> Postnatal maternal anaemia was classified according to the WHO threshold of <12g/dL for women of reproductive age. Severity classifications were defined as mild (11-11.9g/dL), moderate (8-10.9g/dL), and severe (<8g/dL).
<sup>c</sup> The anthropometric measurements at timepoint 1 were conducted by trained medical staff on the research team. Child weight, length, and head circumference measurements were converted to z-scores based on age and sex using Anthro software for WAZ, HAZ, and HCZ. Infants were classified as underweight, stunted, or having microcephaly if they had z-scores of less than -2 SDs.
<sup>d</sup> Fisher's exact test result interpreted due to one or more cells having an expected count of less than 5.
<sup>e</sup> Missing values: monthly household income ( $n = 19$ ), maternal employment ( $n = 2$ ), COVID-19 effect on food insecurity ( $n = 7$ ), infant age at study visit 1 ( $n = 52$ ), infant HIV infection ( $n = 165$ ), infant microcephaly at study visit 1 ( $n = 6$ ), infant underweight at study visit 1 ( $n = 6$ ), infant stunting at study visit 1 ( $n = 9$ ).
\* $p$ is significant at $<0.05$ , \*\* $p$ is significant at $<0.01$ , \*\*\* $p$ is significant at $<0.001$ .

There was overlap between samples across timepoints with 85% (204/240) of mothers with haemoglobin data at postnatal study visit 1, also having antenatal data as described in the previous section. Antenatal maternal anaemia status and postnatal maternal anaemia status were significantly positively associated (*n* = 204), *r* = 0.3, *p* <0.001.

#### Child Anaemia

Child haemoglobin was available from FBCs across the first year of life at study visit 1 (*n*=195; Mean [SD] age of 3.79 [0.77] months, age range of 2 – 5.85 months), study visit 2 (*n*=173; Mean [SD] of 8.54 [1.59] months, age range of 5.29 – 12.20 months), and study visit 3 (*n*=99; Mean (SD) age of 14.16 [1.25], age range of 12 – 18.05 months). Of these infants (Table 3), the prevalence of child anaemia was 51.28% (100/195), 27.17% (47/173), and 20.20% (20/99), respectively. Overall, 46.82% (125/267) were classified as anaemic at least once in the first 6-12 months of life (by study visit 2), and 48.10% (136/283) were classified as anaemic at least once in the first 12-18 months of life (by study visit 3). Risk factors for child anaemia in this sample included being underweight (6% of anaemic infants underweight versus 0% of non-anaemic infants at study visit 1), antenatal alcohol exposure (12.77% of anaemic infants versus 2.38% of non-anaemic infants at study visit 2), and antenatal maternal anaemia (55% of anaemic infants versus 35.71% of non-anaemic infants at study visit 3).

**Table 3.**
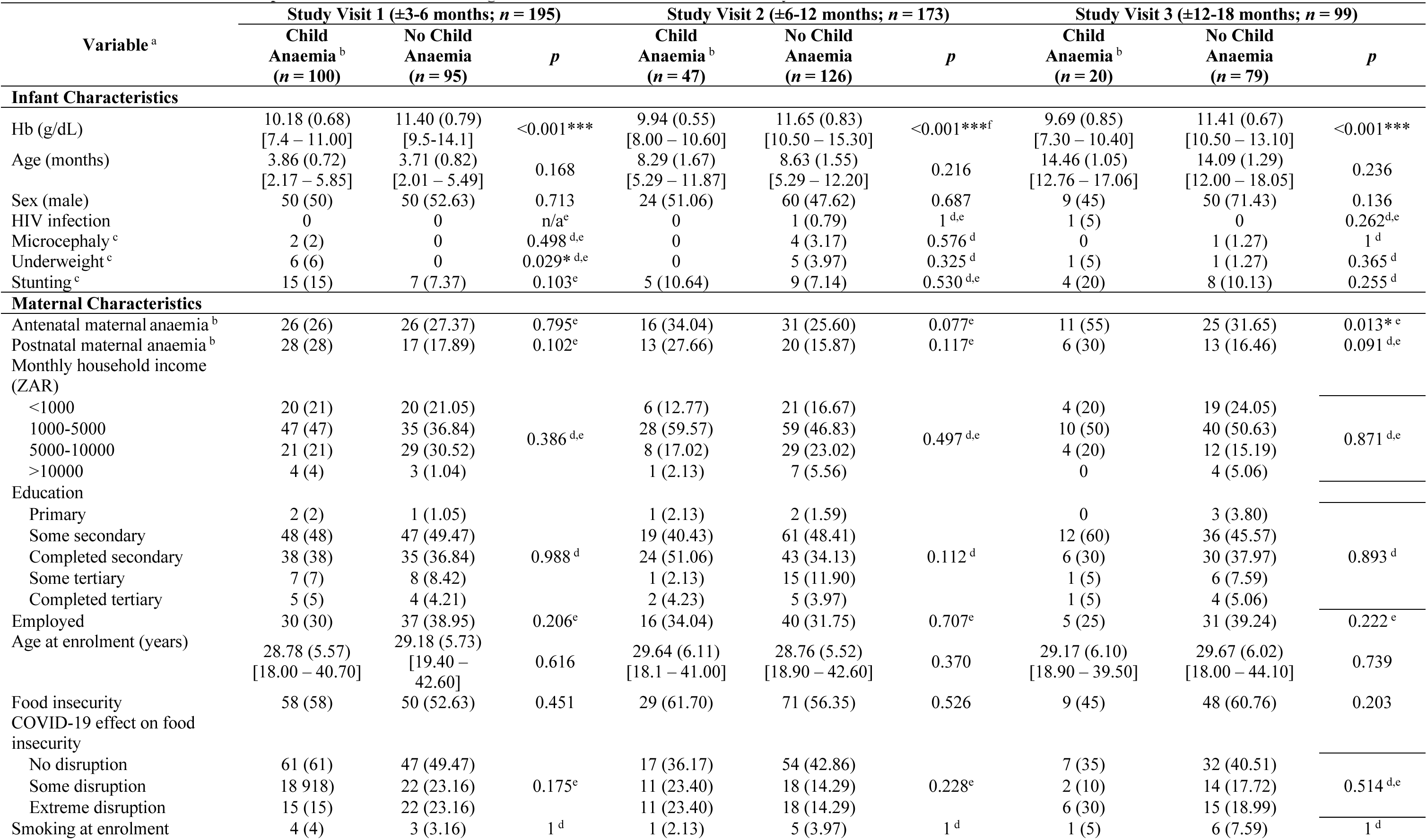
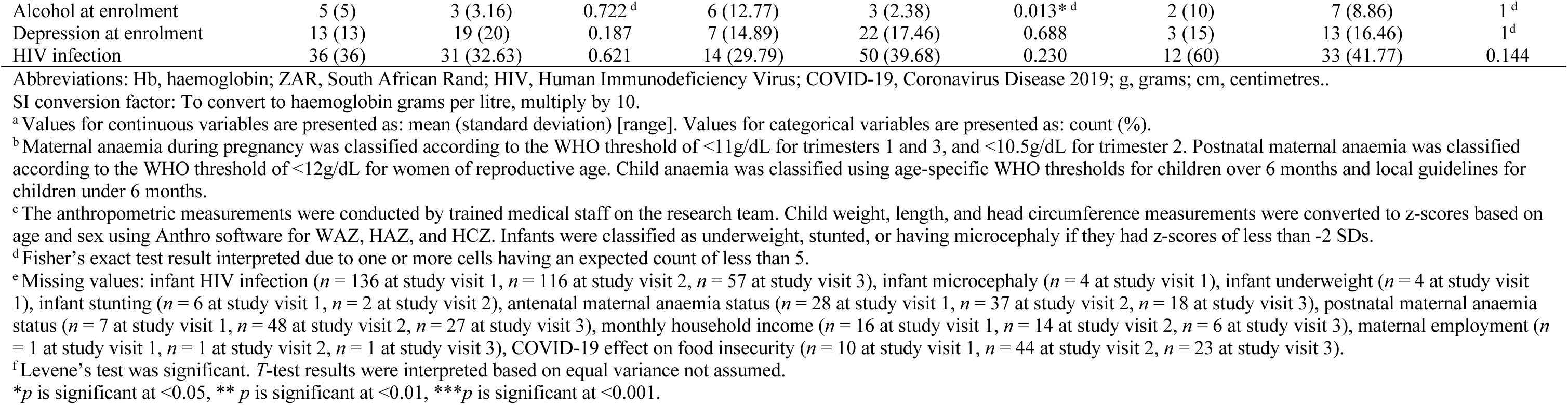
Maternal and Infant Sample Characteristics According to Child Anaemia Status Across Study Visits.

While antenatal maternal anaemia status was not significantly correlated with child anaemia status at visit 1 (*n* = 167, *r*= −0.20, *p*= 0.796) or study visit 2 (*n* = 136, *r* = 0.152, *p* = 0.078), a significantly positive association was observed at study visit 3 (*n* = 81, *r* = 0.277, *p* = 0.012). Postnatal maternal anaemia status was not significantly correlated with child anaemia at study visit 1 (*n* = 188, r = 0.119, p = 0.103), or with child anaemia at later timepoints: study visit 2 (*n* = 125, r = 0.140, p = 0.119), or study visit 3 (*n* = 72, *r* = 0.211, *p* = 0.076).

### Dimension 2: Iron Metrics

Given that iron deficiency is the most common cause of anaemia [1, 33], a detailed description of iron metrics was reported for all mothers and infants with FBCs from any timepoint. The aim of this dimension of the results was to characterise the profile, prevalence, and severity of iron deficiency in the South African context in both anaemic and non-anaemic individuals, while accounting for inflammation using previously established methods from the WHO [23] and BRINDA [24, 25]. In turn, this data was used to determine the extent to which we may be underestimating iron deficiency in this context, and to gain insight into the necessary metrics that may be needed to make a valid diagnosis in high-risk communities.

#### Biomarkers of Inflammation

For all mothers and infants with serum ferritin data available, inflammatory biomarkers were used as indicators of inflammation. As per WHO [23, 46], *hs*CRP and AGP concentrations (descriptive statistics for the full group presented in Supplementary Information Table S2) were used to assess the prevalence of acute (*hs*CRP >5mg/L) and chronic inflammation (AGP >1g/L) status for mothers and infants across study visits (Table 5).

Given that inflammation is known to transiently elevate serum ferritin concentrations [20, 21], the associations between *hs*CRP, AGP, and unadjusted serum ferritin concentrations were explored to assess the trend in this sample. Overall, *hs*CRP and AGP concentrations were positively correlated with unadjusted serum ferritin concentrations for mothers and infants across study visits (Table 4). This was found to be significant for *hs*CRP in mothers postnatally and infants at study visit 2, and AGP in mothers antenatally and postpartum, and in infants postnatally at study visit 2 and study visit 3.

**Table 4.** Associations between Inflammatory Biomarker Concentrations and Unadjusted Serum Ferritin Concentrations In Mothers and Infants Across Study Visits.

| Inflammatory Biomarker | Unadjusted Serum Ferritin |  |  |  |  |
| --- | --- | --- | --- | --- | --- |
|  | Antenatal Maternal <sup>a</sup><br>(*n* = 109) | Postnatal Maternal <sup>a</sup><br>(*n* = 234) | Child Visit 1 <sup>a</sup><br>(*n* = 196) | Child Visit 2 <sup>a</sup><br>(*n* = 158) | Child Visit 3 <sup>a</sup><br>(*n* = 96) |
| *hsCRP* | 0.117 | 0.400*** | 0.076 | 0.188* | 0.057 |
| *AGP* | 0.453*** | 0.367*** | 0.025 | 0.294*** | 0.371*** |

Abbreviations. *hsCRP* = highly sensitive C-Reactive Protein, *AGP* = Alpha-1Acid Glycoprotein
<sup>a</sup> Missing values: Antenatal Maternal *hsCRP* (*n* = 4), postnatal maternal *hsCRP* (*n* = 10), infant *hsCRP* (*n* = 6 at study visit 1, *n* = 5 at study visit 3), infant *AGP* (*n* = 3 at study visit 1, *n* = 2 at study visit 2, *n* = 3 at study visit 3 )
\**p* is significant at <0.05, \*\**p* is significant at <0.01, \*\*\**p* is significant at <0.001

#### Antenatal Maternal Iron Deficiency

In this study (Table 5), a sample of 109 mothers (Mean (SD) age at enrolment: 28.75 (5.75) years; range: 19 – 40.4 years) had antenatal serum ferritin available from Khula enrolment in pregnancy. The prevalence of antenatal maternal iron deficiency (serum ferritin > 15μg/L) using unadjusted serum ferritin concentrations was 18.35% (20/109). However, adjusting for inflammation using Methods A and B increased the prevalence to 55.04% (60/109; 36.69 percentage point increase) and 38.53% (42/109; 20.18 percentage point increase), respectively (Figure 2). Adjusting sTfR concentrations using the BRINDA approach made no difference to the estimates of iron-deficient erythropoiesis (high sTfR) due to very few (*n* = 4) of the mothers having raised AGP.

**Table 5.**
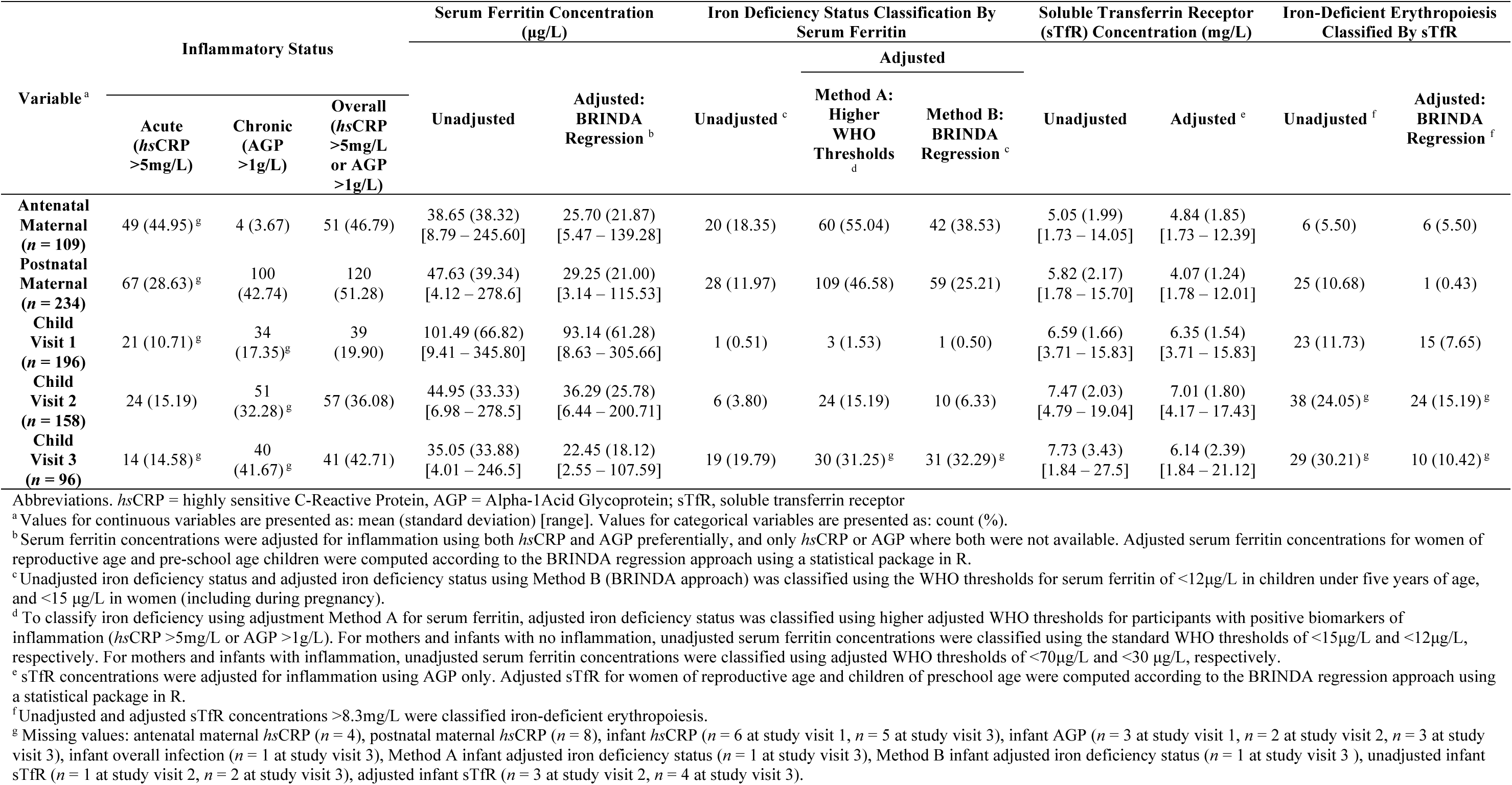
Estimated Iron Deficiency and Iron-Deficient Erythropoiesis Status of Mothers and Infants With and Without Adjustment for Inflammation Across Study Visits.

**Figure 2.**
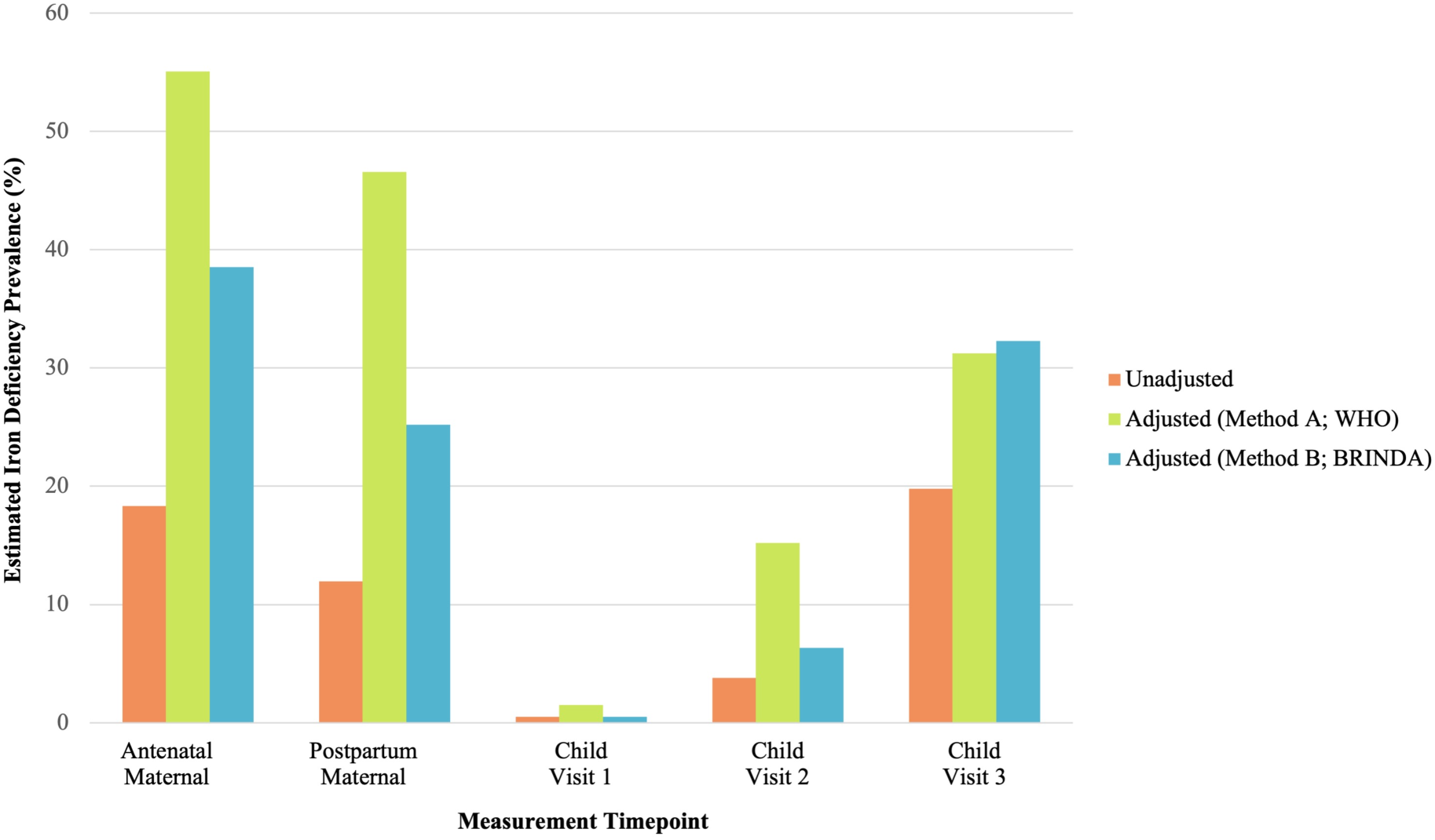
Estimated Prevalence of Iron Deficiency After Adjustment for Inflammation Using Two Methods in Mothers and Infants Across Study Visits

#### Postnatal Maternal Iron Deficiency

At the first study visit 1 (approximately 3-6 months postpartum), 234 mothers (Mean (SD) age at study visit 1: 28.94 (5.58) years; range: 18 – 44.1 years) had serum ferritin available from postnatal FBCs (Table 5). The prevalence of postnatal maternal iron deficiency (serum ferritin > 15μg/L) using unadjusted serum ferritin concentrations was 11.97% (28/234). However, adjusting for inflammation using Methods A and B increased the prevalence to 46.58% (109/234; 34.61 percentage point increase) and 25.21% (59/234; 13.24 percentage point increase), respectively (Figure 2). Adjusting sTfR concentrations based on AGP using the BRINDA approach decreased the estimated prevalence of iron-deficient erythropoiesis from 10.68% (25/234) to 0.43% (1/234; 10.25 percentage point decrease).

Overall, iron deficiency prevalence was consistently lower postpartum than in pregnancy, based on unadjusted and adjusted estimates. However, the prevalence of iron deficiency in mothers, both during and after pregnancy, was underestimated when inflammatory biomarkers (*hs*CRP and AGP) were unaccounted for (Figure 2). In contrast, the estimated prevalence of iron-deficient erythropoiesis was overestimated in postpartum mothers with chronic inflammation (raised AGP).

#### Child Iron Deficiency

At study visit 1, 196 infants (Mean (SD) age: 3.77 (0.77) months; range: 2 – 5.85 months) had serum ferritin available (Table 5). The prevalence of child iron deficiency (serum ferritin > 12μg/L) using unadjusted serum ferritin concentrations was 0.51% (1/196). Adjustment for inflammation using Method A only marginally increased the prevalence to 1.53% (3/196; 1.02 percentage point increase). No change in prevalence was observed using adjustment Method B (Figure 2). Adjusting sTfR concentrations based on AGP using the BRINDA approach decreased the estimated prevalence of iron-deficient erythropoiesis from 11.73% (23/196) to 7.65% (15/196; 4.08 percentage point decrease).

At study visit 2, 158 infants (Mean (SD) age: 8.53 (1.59) months; range: 5.29 – 12.20 months) had serum ferritin available (Table 5). The prevalence of child iron deficiency (serum ferritin > 12μg/L) using unadjusted serum ferritin concentrations was 3.80% (6/158). Adjustment for inflammation using Methods A and B marginally increased the prevalence to 15.19% (24/158; 11.39 percentage point increase) and 6.33% (10/158; 2.53 percentage point increase), respectively (Figure 2). Adjusting sTfR concentrations based on AGP using the BRINDA approach decreased the estimated prevalence of iron-deficient erythropoiesis from 24.05% (38/158) to 15.19% (24/158; 8.86 percentage point decrease).

At study visit 3, 96 infants (Mean (SD) age: 14.17 (1.26) months; range: 12 – 18.05 months) had serum ferritin available (Table 5). The prevalence of child iron deficiency (serum ferritin > 12μg/L) using unadjusted serum ferritin concentrations was 19.79% (19/96). Adjustment for inflammation increased the prevalence of estimated iron deficiency to 31.25% (30/96; 11.46 percentage point increase) and 32.29% (31/96; 12.5% increase) for Methods A and B, respectively (Figure 2). Adjusting sTfR concentrations based on AGP using the BRINDA approach decreased the estimated prevalence of iron-deficient erythropoiesis from 30.21% (29/96) to 10.42% (10/96; 19.79 percentage point decrease).

Overall, the prevalence of iron deficiency in childhood was relatively low at study visits 1 and 2, but increased with age across the first 12-18 months of life. This trajectory was more prominent once inflammation (*hs*CRP and AGP) had been accounted for, with unadjusted serum ferritin underestimating the prevalence of child iron deficiency (Figure 2). Similarly, the estimated prevalence of iron-deficient erythropoiesis was consistently overestimated when not adjusted for chronic inflammation (AGP), particularly by study visit 3.

#### Sample Characteristics by Iron Deficiency Status

Sample characteristics across groups for mothers and infants were reported based on iron deficiency classification with adjustment for inflammation using BRINDA, which is likely to be more specific (Supplementary Information, Tables S3, S4, S5). Group differences were not assessed for infants at study visit 1 given that only one child was found to be iron deficient at this timepoint. Overall, after accounting for inflammation, adjusted serum ferritin concentrations were significantly lower in mothers and infants with iron deficiency across timepoints. Similarly, adjusted sTfR concentrations were significantly higher, suggestive of iron-deficient erythropoiesis, in mothers and infants with iron deficiency across timepoints. Maternal and infant characteristics were similar between groups.

### Dimension 3: Iron Deficiency Anaemia Metrics

#### Iron Metrics

To gain further insight into the prevalence of IDA, this was estimated in a sub-group of anaemic mothers and infants with adjusted serum ferritin data and MCV data for comparison, as detailed below. The prevalence of IDA was reported using anaemia classifications and adjusted iron deficiency classifications from the BRINDA approach (Method B; Table 6).

**Table 6.**
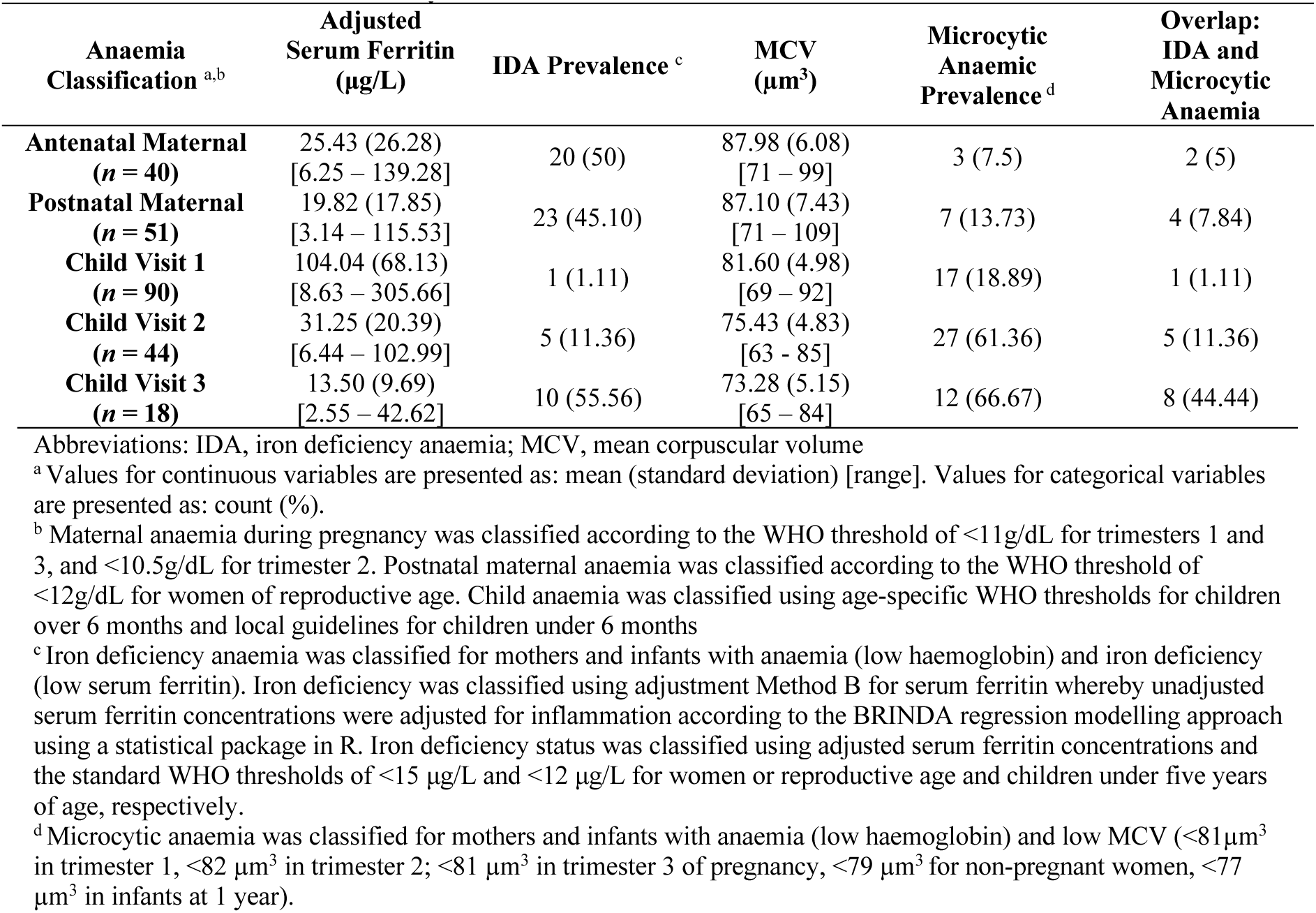
Iron Deficiency and Microcytic Anaemia Classification Using Adjusted Serum Ferritin and MCV Data for Mothers and Infants Across Study Visits.

| Anaemia Classification <sup>a,b</sup> | Adjusted Serum Ferritin (µg/L) | IDA Prevalence <sup>c</sup> | MCV (µm<sup>3</sup>) | Microcytic Anaemic Prevalence <sup>d</sup> | Overlap: IDA and Microcytic Anaemia |
| --- | --- | --- | --- | --- | --- |
| **Antenatal Maternal**<br>(n = 40) | 25.43 (26.28)<br>[6.25 – 139.28] | 20 (50) | 87.98 (6.08)<br>[71 – 99] | 3 (7.5) | 2 (5) |
| **Postnatal Maternal**<br>(n = 51) | 19.82 (17.85)<br>[3.14 – 115.53] | 23 (45.10) | 87.10 (7.43)<br>[71 – 109] | 7 (13.73) | 4 (7.84) |
| **Child Visit 1**<br>(n = 90) | 104.04 (68.13)<br>[8.63 – 305.66] | 1 (1.11) | 81.60 (4.98)<br>[69 – 92] | 17 (18.89) | 1 (1.11) |
| **Child Visit 2**<br>(n = 44) | 31.25 (20.39)<br>[6.44 – 102.99] | 5 (11.36) | 75.43 (4.83)<br>[63 – 85] | 27 (61.36) | 5 (11.36) |
| **Child Visit 3**<br>(n = 18) | 13.50 (9.69)<br>[2.55 – 42.62] | 10 (55.56) | 73.28 (5.15)<br>[65 – 84] | 12 (66.67) | 8 (44.44) |

Abbreviations: IDA, iron deficiency anaemia; MCV, mean corpuscular volume
<sup>a</sup> Values for continuous variables are presented as: mean (standard deviation) [range]. Values for categorical variables are presented as: count (%).
<sup>b</sup> Maternal anaemia during pregnancy was classified according to the WHO threshold of <11g/dL for trimesters 1 and 3, and <10.5g/dL for trimester 2. Postnatal maternal anaemia was classified according to the WHO threshold of <12g/dL for women of reproductive age. Child anaemia was classified using age-specific WHO thresholds for children over 6 months and local guidelines for children under 6 months
<sup>c</sup> Iron deficiency anaemia was classified for mothers and infants with anaemia (low haemoglobin) and iron deficiency (low serum ferritin). Iron deficiency was classified using adjustment Method B for serum ferritin whereby unadjusted serum ferritin concentrations were adjusted for inflammation according to the BRINDA regression modelling approach using a statistical package in R. Iron deficiency status was classified using adjusted serum ferritin concentrations and the standard WHO thresholds of <15 µg/L and <12 µg/L for women or reproductive age and children under five years of age, respectively.
<sup>d</sup> Microcytic anaemia was classified for mothers and infants with anaemia (low haemoglobin) and low MCV (<81µm<sup>3</sup> in trimester 1, <82 µm<sup>3</sup> in trimester 2; <81 µm<sup>3</sup> in trimester 3 of pregnancy, <79 µm<sup>3</sup> for non-pregnant women, <77 µm<sup>3</sup> in infants at 1 year).

Iron deficiency was found to account for approximately half of anaemia cases (Table 6) in mothers antenatally (50%; 20/40) and postnatally (45.10%; 23/51), and in infants at 12-18 months postnatally (55.56%; 10/18). The prevalence of IDA in infants at 3-6 (1.11%; 1/90) months and 6-12 (11.36%; 5/44) months was low, indicating that anaemia at these timepoints may not be due to iron deficiency.

#### MCV Metrics

Given the widespread clinical use of MCV as an indicator of anaemia aetiology with microcytic anaemia recognised as a proxy for iron deficiency, this classification was reported alongside iron metrics for comparative purposes. In a sample of anaemic mothers and infants with both adjusted serum ferritin and MCV data, the prevalence of microcytic anaemia was reported to assess its validity as a marker of iron deficiency relative to current reference standards using iron metrics (Figure 3).

**Figure 3.**
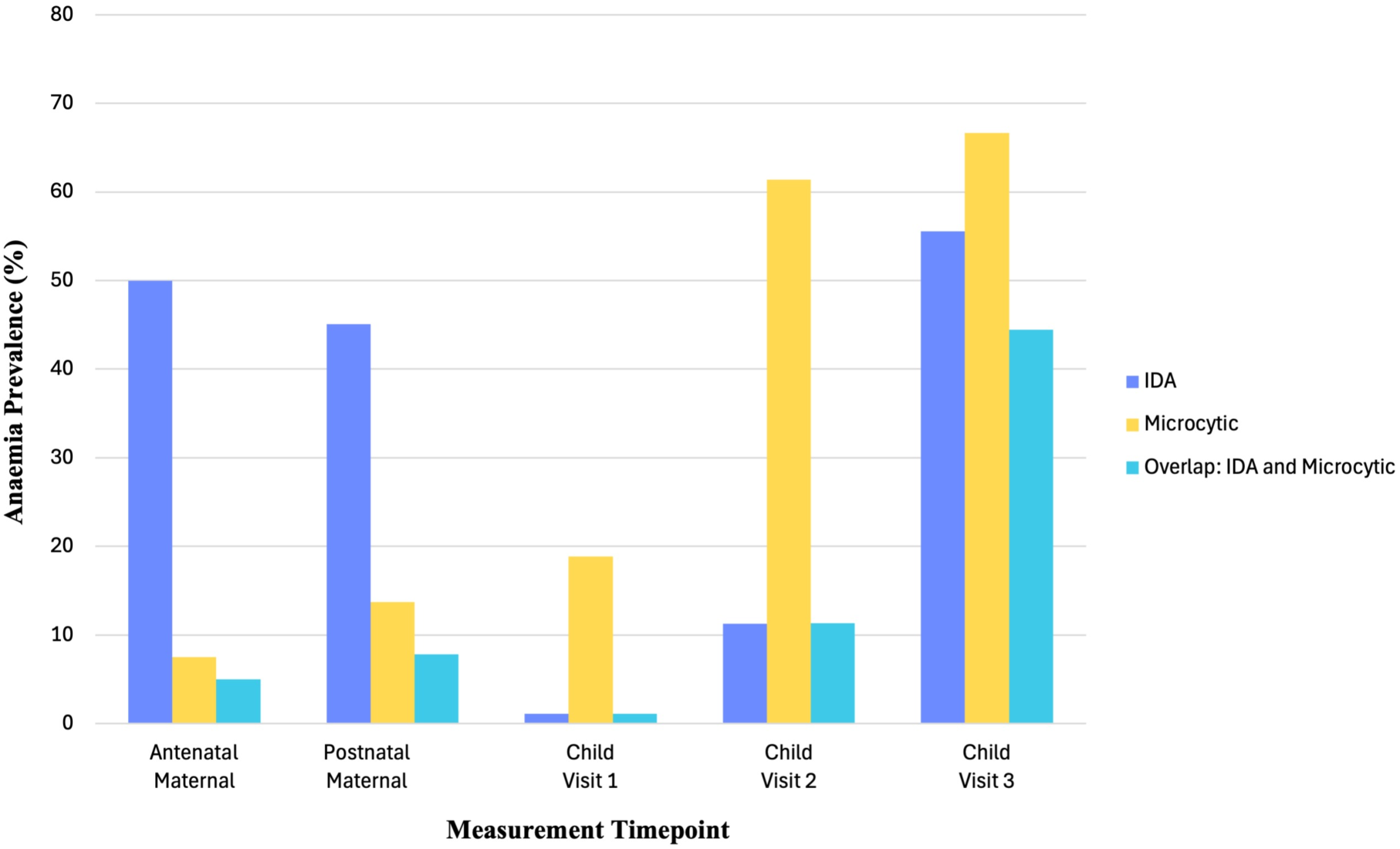
Relative Prevalence of Iron Deficiency Anaemia, Microcytic Anaemia, and Their Overlap in Mothers and Infants Across Study Visits

In comparison to actual IDA classification estimates based on iron metrics (Table 6), microcytic anaemia was found to underestimate iron deficiency in pregnant and postpartum mothers (7.5% microcytic anaemia versus 50% IDA in pregnancy; 13.73% microcytic anaemia versus 45.10% IDA postpartum) and overestimate IDA in infants at 3-6 (18.89% microcytic anaemia versus 1.11% IDA) and 6-12 months (61.36% microcytic anaemia versus 11.36% IDA). Furthermore, there was poor overlap between IDA and microcytic anaemia classifications in pregnant and postpartum women, and in the first year of infancy (Figure 3). Of the pregnant and postpartum mothers with microcytic anaemia, only 66.67% (2/3) and 57.14% (4/7) had IDA, respectively. Similarly, of the infants at 3-6 months and 6-12 months with microcytic anaemia, only 5.88% (1/17) and 18.52% (5/27) had IDA, respectively.

However, the estimated prevalence of IDA and microcytic anaemia was similar in infants at 12-18 months (66.67% microcytic anaemia versus 55.56% IDA) and there was more overlap with 66.67% (8/12) of infants with microcytic anaemia also having IDA. This suggests that microcytic anaemia may be a better proxy for iron deficiency at this age.

## DISCUSSION

This report is one of the first to characterise the prevalence and profile of anaemia, iron deficiency, and IDA with adjustment for inflammation in pregnant and postpartum women, as well as their infants in South Africa. By comparing unadjusted and adjusted estimates of iron deficiency using two different approaches (WHO [23] versus BRINDA [24]), it reveals the extent to which this health concern may be underestimated in countries with a high burden of infection.

### Maternal IDA

In this birth cohort, the prevalence of anaemia in pregnancy was found to be 34.74% with most mothers having mild or moderate anaemia. This is similar to previous estimates for LMICs [1] and research from other high-risk South African birth cohorts, corroborating the burden of anaemia in this context [9, 10]. While reported food insecurity was high with exacerbated effects due to COVID-19, there were no group differences in access to food resources, or anthropometric child birth outcomes between anaemic and non-anaemic mothers. This suggests that anaemia in this sample is more likely to be associated with a micronutrient deficiency such as iron rather than gross malnutrition. In considering associated risk factors for anaemia, it is noted that the prevalence of smoking and alcohol use in pregnancy was low across the cohort. However, mothers within the antenatal maternal anaemia group had a higher prevalence of HIV infection, suggesting that inflammation may be an important aetiological factor for consideration in this context.

For mothers with iron and inflammatory metrics in pregnancy, almost half (46.79%) had evidence of inflammation, most of which was acute (raised *hs*CRP). The lower prevalence of biomarkers for chronic inflammation (raised AGP) may be associated with immune modulation typical of the physiology of pregnancy [52, 53]. However, as expected, AGP concentrations were positively correlated with serum ferritin levels, with a similar trend in *hs*CRP. This is consistent with serum ferritin being a well-documented acute-phase reactant that becomes elevated in inflammatory states [20, 21]. In our cohort, while the unadjusted prevalence of iron deficiency was estimated to be 18.35%, adjustment for inflammation using Method A (WHO; higher serum ferritin thresholds) and Method B (BRINDA; regression correction) increased the estimated prevalence to 55.04% (36.69% increase) and 38.53% (20.18% increase), respectively. Taken together, these findings emphasize the importance of adjusting serum ferritin for inflammation during pregnancy to avoid the underestimation of antenatal maternal iron deficiency. Using adjusted estimates of iron deficiency based on BRINDA, it was found that half of maternal anaemia cases in pregnancy could be attributed to iron deficiency (IDA). In contrast, of all antenatal maternal anaemia cases, only 7.5% were classified as microcytic and, of these mothers, only 66.67% also had IDA. This suggests that MCV may not be as valid a proxy for iron deficiency during pregnancy when the burden of inflammation is high.

Mothers with anaemia in pregnancy had a higher risk of having anaemia postpartum, compared to those who were not anaemic in pregnancy. While the prevalence of postnatal maternal anaemia was still high (22.50%), it was lower than antenatal maternal anaemia; this was expected given that pregnancy represents a period of heightened metabolic demand [16, 54, 55]. However, the risk profile remained similar, with the only group difference being a higher proportion of anaemic mothers also living with HIV.

The prevalence of postnatal maternal inflammation remained high (51.28%), with evidence for both acute (raised *hs*CRP) and chronic inflammation (raised AGP). Both inflammatory biomarker concentrations were separately associated with serum ferritin levels, confirming that inflammation contributes to elevated serum ferritin levels. The estimated prevalence of iron deficiency was lower postpartum than during pregnancy, but increased from 11.97% to 46.58% (34.61 % increase) and 25.21% (13.24 % increase) when adjusting for inflammation using Methods A (WHO; higher serum ferritin thresholds) and B (BRINDA; regression correction), respectively. Overall, this evidence suggests that, where inflammation is not accounted for, the prevalence of iron deficiency may be underestimated while the prevalence of iron-deficient erythropoiesis may be overestimated in postpartum women. While microcytic anaemia classification using traditional approaches suggested that only 13.73% of postnatal anaemia cases were likely to be IDA, BRINDA-adjusted iron metrics demonstrated that 45.10% of anaemia cases were in fact attributable to iron deficiency. Furthermore, only 57.14% of mothers with microcytic anaemia also had IDA. Therefore, in our cohort, MCV remained a poor proxy for IDA in early postpartum mothers with active inflammation. These results suggest the prevalence of maternal anaemia in this South African community is high, particularly during pregnancy, with most cases being associated with iron deficiency.

Despite the high burden of anaemia, iron deficiency, and IDA, very few mothers had an accurate understanding of their own anaemia status at enrolment, with only four mothers (3.74%) in the anaemic group reporting their own status accurately. This highlights the need for public engagement and awareness around the importance of nutrition, supplementation, and management of conditions including iron deficiency, substance use, HIV, and other infections with an increased focus on pregnancy as a priority [3, 56].

### Child IDA

The prevalence of child anaemia in this cohort was also high with 46.82% and 48.10% of infants being classified as anaemic at least once by the second and third study visit at approximate ages of 6-12 months and 12-18 months, respectively. Most cases of child anaemia seemed to be evident early on with the prevalence of new diagnoses decreasing from 51.28% to 20.20% over the 3 postnatal study visits. This is consistent with research from Kenya and Togo suggesting that the risk of anaemia is greatest within the first year of life, progressively decreasing thereafter [57, 58]. It is suggested that this pattern may be due to a rapid decline in haemoglobin and a change in globin chains immediately after birth, when the hypoxic uterine environment is exchanged for oxygen-rich air exposure [3].

In this study, early risk factors included malnutrition, as indicated by underweight classification, and prenatal alcohol exposure. Similarly, infants whose mothers were anaemic during pregnancy were more likely to be anaemic themselves by 12-18 months of age. This corroborates research from across multiple African countries, identifying antenatal maternal anaemia as an important predictor of childhood anaemia [58–60]. While child exposure to maternal HIV in utero was high, very few infants were found to have HIV, most likely due to good maternal access to effective ART in pregnancy for this cohort [61]. However, there was still evidence for inflammation amongst the infants in early life, with approximately 20%-43% of infants having positive biomarkers (*hs*CRP >5mg/L or AGP >1g/L), particularly for chronic inflammation (AGP >1g/L). While there was no consistent relationship between acute or chronic inflammation and iron deficiency within the first 3-6 months of life in our sample, significant positive associations between inflammatory biomarkers (*hs*CRP and AGP) and serum ferritin concentrations emerged after 6 months of age.

Based on unadjusted serum ferritin, a low estimated prevalence of iron deficiency was observed within the first 3-6 and 6-12 months (study visits 1 and 2), and adjustment for inflammation only marginally increased this. Furthermore, very few anaemia cases were attributed to iron deficiency in this early period (1.11% and 11.26%), despite the high proportion of anaemic infants being classified as having microcytic anaemia (18.89% and 61.36%). This may be due to the protective effects of foetal iron loading in pregnancy, with maternal iron stores typically sustaining iron demands in infants for the first 6 months when they are unable to acquire and absorb much iron from their diet [16, 54, 55]. Additionally, exclusive breastfeeding may have protective effects within the first 4-6 months of infancy due to the high bioavailability of iron in breastmilk (despite low iron concentrations) [3].

Overall, the biological mechanisms for anaemia in early life in this cohort are unclear with unstable associations between inflammatory biomarkers and iron metrics. This is most likely due to the fact that metabolic and immune processes are immature in infancy [62], with transitions between food sources from immune-protective breastmilk [63, 64] to solid foods typically occurring in parallel [65]. Early infancy is a period of great flux, given the increased risk of severe infections, the high prevalence of environmental exposures, and maturing physiological processes. Therefore, stable patterns or relationships between factors may be harder to disentangle during this 3–12-month window. It is also important to note that MCV may be a poor proxy for IDA at this age given that only 5.88% and 18.52% of infants with microcytic anaemia were classified with IDA at 3-6 and 6-12 months, respectively.

By 12-18 months (study visit 3), iron deficiency anaemia patterns in our child cohort stabilised, with prevalence and clinical associations becoming more similar to their mothers. At this timepoint, iron deficiency was more prevalent than earlier in infancy. This is consistent with research suggesting that the risk of iron deficiency is greatest between 6 and 24 months when maternal iron sources are depleted and infant growth is exponential [3]. The adjustment for inflammation using both Method A (WHO) [23] and Method B (BRINDA) [24] increased the estimated prevalence of iron deficiency from 19.71% to 31.25% (11.54% increase) and 32.29% (12.5% increase), respectively. Furthermore, the estimated prevalence of iron deficient erythropoiesis was overestimated by 19.79% when not adjusted for chronic inflammation (AGP). As seen in mothers antenatally and postnatally, iron deficiency (based on inflammation-adjusted values) accounted for approximately 50% of anaemia cases. Unlike previous maternal and child timepoints, MCV seems to be a more reliable proxy for iron deficiency in infants at 12-18 months with similar prevalence estimates for microcytic anaemia (66.67%) and IDA (55.56%), and more overlap (66.67% of infants with microcytic anaemia being classified as having IDA). The positive association between antenatal maternal and child anaemia at this timepoint, provides a firmer bases for aetiological comparisons.

The complex interplay between overlapping risk factors for childhood anaemia in Africa [57–60, 66–68] highlights the importance of inflammatory biomarkers to support an improved understanding of risk and to inform strategies for intervention at an individual and population level. This is particularly relevant at 12-18 months when the prevalence of IDA is likely to be underestimated in infants with active inflammation. However, further research on inflammation and the immune role in IDA in infants may provide further insight into this evolving relationship, particularly in the first year of life.

### Considerations for Interpreting IDA

Given the results of this study, it is clear that inflammation is an important factor for consideration in the assessment of anaemia, iron deficiency, and their overlap. However, due to limited evidence for the epidemiology of iron deficiency and IDA in Africa, where infection is prominent, more information is needed on valid assessment approaches and the interpretation of iron metrics, particularly in pregnancy and early infancy.

As shown in illustrative Figure 4, previously recognised risk factors for anaemia include food insecurity and malnutrition [3, 18, 19], infectious diseases such as HIV and comorbidities [3, 17, 69], exposure to toxins such as alcohol and smoking that affect biological/haematological processes [70, 71] and nutritional behaviours [3, 72], inadequate access to healthcare services, adequate sanitation, and fortified foods [4], and limited public awareness and knowledge of nutrition and anaemia [73, 74]. It is well-known that many of these drivers of anaemia aetiology [2] are more prevalent in LMICs where there are high levels of poverty, low levels of education, limited healthcare resources, and increased prevalence of infectious diseases [1, 6, 7, 33]. Classifying anaemia is based on WHO haemoglobin thresholds with age-specific and trimester-specific cutoffs for infants and women, respectively [41].

**Figure 4.**
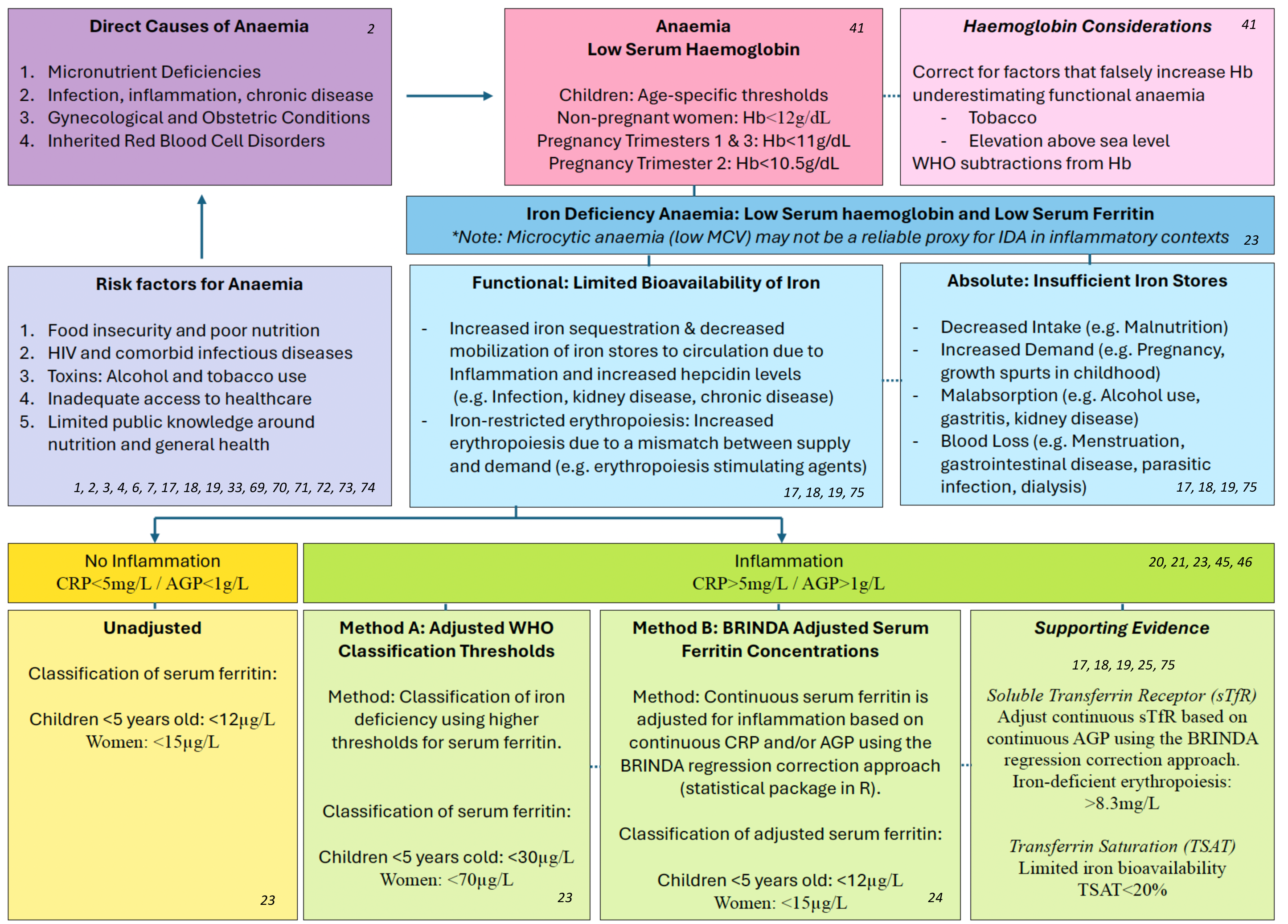
Considerations for Understanding and Interpreting Iron Deficiency Anaemia with Inflammation in Pregnancy and Infancy Abbreviations: Hb, haemoglobin, IDA, iron Deficiency Anaemia, MCV, Mean Corpuscular Volume, highly sensitive C-Reactive Protein, AGP = Alpha-1AcidGlycoprotein; sTfR, soluble transferrin receptor

However, smoking and high altitudes require special consideration in haemoglobin measurement as they may transiently elevate haemoglobin levels resulting in the underestimation of functional anaemia. The WHO guidelines include recommended subtractions to correct haemoglobin measurements based on the number of cigarettes smoked per day and the degree of elevation above sea level [41].

As described earlier, overlapping risk factors and mechanisms may contribute to absolute (limited iron stores) or functional (limited iron bioavailability) iron deficiency in LMICs [17–19, 75]. For example, malnutrition (decreased intake) during childhood or pregnancy (periods of increased demand) may be further exacerbated by gastritis or alcohol consumption, both of which contribute to the malabsorption of iron. In parallel to these drivers of absolute iron deficiency, there may be a functional overlay associated with inflammation. Typically, inflammation promotes the expression of hepcidin, resulting in increased iron sequestration and decreased mobilisation of iron stores into circulation for use (limited bioavailability).

Given that serum ferritin (WHO metric for classifying iron deficiency) [23] is an acute-phase reactant, it becomes transiently elevated in response to inflammation [20, 21]. Therefore, inflammatory biomarkers of acute (*hs*CRP >5mg/L) or chronic inflammation (AGP >1g/L) [45] are important for the valid interpretation of iron metrics when the burden of infectious disease is high [23, 46]. This was an important factor for consideration in this study.

Based on our results, we highlight two available methods for interpreting iron metrics with adjustment for inflammation. Method A uses adjusted WHO thresholds to classify iron deficiency in cases with inflammation [23]. Method B uses the BRINDA regression approach to corrects serum ferritin based on *hs*CRP and AGP [24]. While Method A [23] is likely to be more sensitive, it is a less specific measure of iron deficiency due to its generalised approach. In comparison, Method B [24] is expected to have higher specificity as it corrects serum ferritin on an individual level, but the model is still being refined. As demonstrated in this research, additional supporting evidence may be derived from sTfR which serves as an indicator of iron-deficient erythropoiesis [25], but requires adjustment for inflammation BRINDA [25]. While transferrin saturation (TSAT) was unavailable for inclusion in this study, it has also been identified as a reliable proxy for iron bioavailability [17–19, 75].

## Conclusion and Future Directions

This study is one of the first to characterise IDA in South African mothers (during and after pregnancy) and infants with adjustment for inflammation. Given the high burden of infection such as HIV in this country, the results provide novel insight into the impact of inflammation on haemoglobin and iron metrics. This paper demonstrates the extent to which iron deficiency and IDA may be underestimated if inflammation is not accounted for in populations where infectious burden is high. Our findings suggests that microcytic anaemia (based on MCV) may be a less useful proxy for IDA in pregnant and postpartum mothers or infants under 1 year of age within the context of inflammation. We highlight the value of adjusting serum ferritin interpretation using inflammatory biomarkers (*hs*CRP and AGP). The complex aetiology of anaemia is emphasized with informed considerations for understanding overlapping risk factors in this critical period. Given the high prevalence of IDA and the even higher prevalence of iron deficiency, valid interpretation of iron metrics is key for improved screening, context-specific prevention efforts, and optimised interventions strategies around iron supplementation and management of infectious disease [3, 56].

The refinement of population-specific reference ranges is particularly necessary in communities of African descent, where it is estimated that genetic differences may translate into naturally lower haemoglobin concentrations [3]. While correction methods for inflammation adjustment are useful tools for detecting iron deficiency in countries with a high burden of infection, they are still based on existing reference ranges which may not be appropriate. Therefore, ongoing efforts to reduce IDA should include epidemiological data from LMICS (including pregnancy data) for representation across contexts with consideration of inflammation as a key aetiological contributor. Lastly, more research is required to understand the complex and evolving role of the immune system in IDA during early life.

Future work, such as the Gates Foundation funded ReMAPP (Redefining Maternal Anemia in Pregnancy and Postpartum) study, focused on identifying specific thresholds for anaemia and iron deficiency in pregnancy and infancy at a larger scale across LMICS, will be important in establishing appropriate thresholds for these periods. In turn, this work may inform current intervention research including the Randomised controlled trial of the Effect of Intra-Venous Iron on Anaemia in Malawian Pregnant women (REVAMP; Gates Foundation) [76]. This multifactorial approach to understanding, preventing, and treating anaemia is important for ongoing efforts aimed at improving neurodevelopmental outcomes, particularly in LMICs where many infants are not reaching their full potential [8].

## ABBREVIATIONS

AEQ: Alcohol Exposure Questionnaire

AGP: Alpha-1Acid Glycoprotein

ASSIST: Alcohol, Smoking, and Substance Involvement Screening Test

BRINDA: Biomarkers Reflecting Inflammation and Nutritional Determinants of Anaemia

CDC: Centers for Disease Control

COPE-IS: Coronavirus Perinatal Experiences – Impact Survey

COVID-19: Coronavirus Disease 2019

EPDS: Edinburgh Postnatal Depression Scale

FBC: Full Blood Count

HAZ: Height-for-Age

Hb: Haemoglobin

HCZ: Head-Circumference-for-Age

HIV: Human Immunodeficiency Virus

*hs*CRP: Highly Sensitive C-Reactive Protein

IDA: Iron Deficiency Anaemia

LMIC: Low- and Middle-Income Country

MCV: Mean Corpuscular Volume

NHLS: National Health Laboratory Services

PHDC: Provincial Health Data Centre

ReMAPP: Redefining Maternal Anemia in Pregnancy and Postpartum

REVAMP: Randomised controlled trial of the Effect of Intra-Venous Iron on Anaemia in Malawian Pregnant women

SDG: Sustainable Development Goal

SPSS: Statistical Package for the Social Sciences

sTfR: Soluble Transferrin Receptor

TSAT: Transferrin Saturation

UN: United Nations

UNICEF: United Nations International Emergency Fund

WAZ: Weight-for-Age

WHO: World Health Organization

ZAR: South African Rands

## ARTICLE INFORMATION

### Author Contributions

*Study Conceptualization and Study Design:* J.E. Ringshaw, M.R. Zieff, S. Williams, C.A. Jacobs, D.C. Alexander, M. Gladstone, V. Klepac-Ceraj, L.J. Gabard-Durnam, D. Amso, W.P. Fifer, D.K. Jones, D.J. Stein, S.C.R. Williams, K.A. Donald

*Data Collection:* S. Williams, C.A. Jacobs, Khula South African Data Collection Team*

*Data Curation:* J.E. Ringshaw, M.R. Zieff, Z. Goolam Nabi, T. Mazubane

*Data Analysis:* J.E. Ringshaw, M.R. Zieff

*Data Interpretation:* J.E. Ringshaw, M.R. Zieff, S. Williams, C.A. Jacobs, D.C. Alexander, M. Gladstone, V. Klepac-Ceraj, L.J. Gabard-Durnam, D. Amso, W.P. Fifer, D.K. Jones, D.J. Stein, S.C.R. Williams, K.A. Donald

*Writing - Original Draft Preparation:* J.E. Ringshaw, K.A. Donald

*Writing - Critical Review, & Editing:* J.E. Ringshaw, M.R. Zieff, S. Williams, C.A. Jacobs, Z. Goolam Nabi, T. Mazubane, M. Miles, D. Herr, D.C. Alexander, M. Gladstone, V. Klepac-Ceraj, L.J. Gabard-Durnam, D. Amso, W.P. Fifer, D.K. Jones, D.J. Stein, S.C.R. Williams, K.A. Donald

*Administrative, Technical, or Material Support:* M.R. Zieff, S. Williams, C.A. Jacobs, M. Miles, D. Herr

*Supervision:* D.J. Stein, S.C.R. Williams, K.A. Donald

*Khula South African Data Collection Team: Layla Bradford, Simone Williams, Lauren Davel, Tembeka Mhlakwaphalwa, Bokang Methola, Khanyisa Nkubungu, Candice Knipe, Zamazimba Madi, Nwabisa Mlandu

## Data Availability

The de-identified data that support the findings of this study are available upon reasonable request from the corresponding author as per Khula cohort guidelines.

## Acknowledgements

We would like to thank all the mothers and infants who participated in the Khula South Africa birth cohort study and made this research possible. We are also sincerely grateful to all the staff at both the Gugulethu Midwife Obstetric Unit as well as the University of Cape Town Neuroscience Institute who were involved in recruitment and data collection.

## Ethical Considerations

The Khula South Africa birth cohort study (reference: 666/2021) including all analyses on antenatal maternal anaemia received ethical approval from the University of Cape Town Human Ethics Research Committee (HREC). This original research was conducted in partial fulfilment of J.E. Ringshaw’s PhD which also has institutional ethical approval from the University of Cape Town (reference: 782/2022). All mothers provided written informed consent for cohort participation at Khula enrolment, and for their children to participate in study-specific procedures across timepoints. Given the longitudinal nature of the cohort, study consent was obtained from mothers on an annual basis. While specified, approved authors had access to identifiable participant information during data collection, only non-identifiable data was captured and analysed.

## REFERENCES

1. Stevens GA, Paciorek CJ, Flores-Urrutia MC, Borghi E, Namaste S, Wirth JP, et al. National, regional, and global estimates of anaemia by severity in women and children for 2000–19: a pooled analysis of population-representative data. The Lancet Global Health. 2022;10(5):e627–e39.

2. WHO. Accelerating anaemia reduction: a comprehensive framework for action. 2023.

3. Mwangi MN, Mzembe G, Moya E, Verhoef H. Iron deficiency anaemia in sub-Saharan Africa: a review of current evidence and primary care recommendations for high-risk groups. The Lancet Haematology. 2021;8(10):e732–e43.

4. Owais A, Merritt C, Lee C, Bhutta ZA. Anemia among women of reproductive age: an overview of global burden, trends, determinants, and drivers of progress in low- and middle-income countries. Nutrients. 2021;13(8):2745.

5. Mantadakis E, Chatzimichael E, Zikidou P. Iron deficiency anemia in children residing in high and low-income countries: risk factors, prevention, diagnosis and therapy. Mediterranean journal of hematology and infectious diseases. 2020;12(1).

6. Chaparro CM, Suchdev PS. Anemia epidemiology, pathophysiology, and etiology in low-and middle-income countries. Annals of the new York Academy of Sciences. 2019;1450(1):15–31.

7. Hess SY, Owais A, Jefferds MED, Young MF, Cahill A, Rogers LM. Accelerating action to reduce anemia: Review of causes and risk factors and related data needs. Annals of the New York Academy of Sciences. 2023;1523(1):11–23.

8. Donald KA, Wedderburn CJ, Barnett W, Nhapi RT, Rehman AM, Stadler JA, et al. Risk and protective factors for child development: An observational South African birth cohort. PLoS Medicine. 2019;16(9):e1002920.

9. Wedderburn CJ, Ringshaw JE, Donald KA, Joshi SH, Subramoney S, Fouche J-P, et al. Association of Maternal and Child Anemia With Brain Structure in Early Life in South Africa. JAMA Network Open. 2022;5(12):e2244772-e.

10. Ringshaw JE, Hendrikse C, Wedderburn CJ, Bradford LE, Williams SR, Nyakonda CN, et al. Persistent Impact of Antenatal Maternal Anaemia on Child Brain Structure at 6–7 Years of Age: A South African Child Health Study. Research square. 2024.

11. WHO. Comprehensive implementation plan on maternal, infant and young child nutrition. World Health Organization; 2014.

12. WHO. Global nutrition targets 2025: policy brief series 2014 [Available from: https://www.who.int/publications/i/item/WHO-NMH-NHD-14.2.

13. WHO. Trends in maternal mortality 2000 to 2020: estimates by WHO, UNICEF, UNFPA, World Bank Group and UNDESA/Population Division: executive summary. 2023.

14. UN. Global indicator framework for the Sustainable Development Goals and targets of the 2030 Agenda for Sustainable Development 2020 [Available from: https://unstats.un.org/sdgs/indicators/indicators-list/.

15. WHO U. The extension of the 2025 maternal, infant and young child nutrition targets to 2030: WHO. 2021.

16. Georgieff MK. Iron deficiency in pregnancy. American Journal of Obstetrics and Gynecology. 2020.

17. Cappellini M, Musallam K, Taher A. Iron deficiency anaemia revisited. Journal of Internal Medicine. 2020;287(2):153–70.

18. Camaschella C. New insights into iron deficiency and iron deficiency anemia. Blood reviews. 2017;31(4):225–33.

19. Camaschella C. Iron deficiency. Blood, The Journal of the American Society of Hematology. 2019;133(1):30–9.

20. Young MF, Suchdev PS. Effects of inflammation on micronutrient biomarkers associated with anemia. Nutritional Anemia: Springer; 2022. p. 63–73.

21. Tomkins A. Assessing micronutrient status in the presence of inflammation. The Journal of nutrition. 2003;133(5):1649S–55S.

22. Kasvosve I, Gomo ZA, Nathoo KJ, Matibe P, Mudenge B, Loyevsky M, et al. Association of serum transferrin receptor concentration with markers of inflammation in Zimbabwean children. Clinica chimica acta. 2006;371(1-2):130–6.

23. WHO. WHO guideline on use of ferritin concentrations to assess iron status in populations: World Health Organization; 2020.

24. Namaste SM, Ou J, Williams AM, Young MF, Emma XY, Suchdev PS. Adjusting iron and vitamin A status in settings of inflammation: a sensitivity analysis of the Biomarkers Reflecting Inflammation and Nutritional Determinants of Anemia (BRINDA) approach. The American Journal of Clinical Nutrition. 2020;112:458S–67S.

25. Rohner F, Namaste SM, Larson LM, Addo OY, Mei Z, Suchdev PS, et al. Adjusting soluble transferrin receptor concentrations for inflammation: Biomarkers Reflecting Inflammation and Nutritional Determinants of Anemia (BRINDA) project. The American journal of clinical nutrition. 2017;106:372S–82S.

26. BRINDA. Biomarkers Reflecting Inflammation and Nutritional Determinants of Anemia [Available from: https://www.brinda-nutrition.org.

27. Suchdev PS, Namaste SM, Aaron GJ, Raiten DJ, Brown KH, Flores-Ayala R, et al. Overview of the biomarkers reflecting inflammation and nutritional determinants of anemia (BRINDA) project. Advances in Nutrition. 2016;7(2):349–56.

28. Suchdev PS, Williams AM, Mei Z, Flores-Ayala R, Pasricha S-R, Rogers LM, et al. Assessment of iron status in settings of inflammation: challenges and potential approaches. The American journal of clinical nutrition. 2017;106:1626S–33S.

29. Muriuki JM, Mentzer AJ, Webb EL, Morovat A, Kimita W, Ndungu FM, et al. Estimating the burden of iron deficiency among African children. BMC medicine. 2020;18:1–14.

30. Ntenda PA, Chirambo AC, Nkoka O, El-Meidany WM, Goupeyou-Youmsi J. Implication of asymptomatic and clinical Plasmodium falciparum infections on biomarkers of iron status among school-aged children in Malawi. Malaria Journal. 2022;21(1):278.

31. Mwangi MN, Echoka E, Knijff M, Kaduka L, Werema BG, Kinya FM, et al. Iron status of Kenyan pregnant women after adjusting for inflammation using BRINDA regression analysis and other correction methods. Nutrients. 2019;11(2):420.

32. UNAIDS. AIDSinfo [Available from: http://aidsinfo.unaids.org.

33. Stevens GA, Beal T, Mbuya MN, Luo H, Neufeld LM, Addo OY, et al. Micronutrient deficiencies among preschool-aged children and women of reproductive age worldwide: a pooled analysis of individual-level data from population-representative surveys. The Lancet Global Health. 2022;10(11):e1590–e9.

34. Maner BS, Moosavi L. Mean corpuscular volume. 2019.

35. Zieff MR, Miles M, Mbale E, Eastman E, Ginnell L, Williams SC, et al. Characterizing developing executive functions in the first 1000 days in South Africa and Malawi: The Khula Study. Wellcome Open Research. 2024;9(157):157.

36. Humeniuk R, Henry-Edwards S, Ali R, Poznyak V, Monteiro MG, Organization WH. The Alcohol, Smoking and Substance Involvement Screening Test (ASSIST): manual for use in primary care. 2010.

37. WHO. The alcohol, smoking and substance involvement screening test (ASSIST): development, reliability and feasibility. Addiction. 2002;97(9):1183–94.

38. Cox JL, Holden JM, Sagovsky R. Detection of postnatal depression: development of the 10-item Edinburgh Postnatal Depression Scale. The British journal of psychiatry. 1987;150(6):782–6.

39. Kessler RC, Üstün TB. The world mental health (WMH) survey initiative version of the world health organization (WHO) composite international diagnostic interview (CIDI). International journal of methods in psychiatric research. 2004;13(2):93–121.

40. Thomason M, Graham A, VanTieghem M. The cope-is: Coronavirus perinatal experiences–impact survey. 2020.

41. WHO. Guideline on haemoglobin cutoffs to define anaemia in individuals and populations: World Health Organization; 2024.

42. WHO. Haemoglobin Concentrations for the Diagnosis of Anaemia and Assessment of Severity. 2011.

43. Abbassi-Ghanavati M, Greer LG, Cunningham FG. Pregnancy and laboratory studies: a reference table for clinicians. Obstetrics & Gynecology. 2009;114(6):1326–31.

44. CDC. Recommendations to Prevent and Control Iron Deficiency in the United States. 1998.

45. Raiten DJ, Ashour FAS, Ross AC, Meydani SN, Dawson HD, Stephensen CB, et al. Inflammation and nutritional science for programs/policies and interpretation of research evidence (INSPIRE). The Journal of nutrition. 2015;145(5):1039S–108S.

46. WHO. Micronutrient Survey Manual 2020 [Available from: https://iris.who.int/bitstream/handle/10665/336010/9789240012691-eng.pdf?sequence=1.

47. Luo H, Addo, O. Y., Geng, J. BRINDA: Computation of BRINDA Adjusted Micronutrient Biomarkers for Inflammation. R package version 0.1.5 ed 2022.

48. Abraham, Müller, Grüters, Wahn, Schweigert. Minimal inflammation, acute phase response and avoidance of misclassification of vitamin A and iron status in infants– importance of a high-sensitivity C-reactive protein (CRP) assay. International journal for vitamin and nutrition research. 2003;73(6):423–30.

49. Northrop-Clewes CA, Thurnham DI. Biomarkers for the differentiation of anemia and their clinical usefulness. Journal of blood medicine. 2013:11–22.

50. VitMinLab. Measuring the Vit. A and Iron Status [Available from: http://www.nutrisurvey.de/vitmin/.

51. Erhardt JG, Estes JE, Pfeiffer CM, Biesalski HK, Craft NE. Combined measurement of ferritin, soluble transferrin receptor, retinol binding protein, and C-reactive protein by an inexpensive, sensitive, and simple sandwich enzyme-linked immunosorbent assay technique. The Journal of nutrition. 2004;134(11):3127–32.

52. Racicot K, Kwon JY, Aldo P, Silasi M, Mor G. Understanding the complexity of the immune system during pregnancy. American journal of reproductive immunology. 2014;72(2):107–16.

53. Mor G, Cardenas I. The immune system in pregnancy: a unique complexity. American journal of reproductive immunology. 2010;63(6):425–33.

54. Georgieff MK, Ramel SE, Cusick SE. Nutritional influences on brain development. Acta Paediatrica. 2018;107(8):1310–21.

55. Cusick SE, Georgieff MK. The role of nutrition in brain development: the golden opportunity of the “first 1000 days”. The Journal of pediatrics. 2016;175:16–21.

56. McDonald CM, Wessells KR, Stewart CP, Dewey KG, de Pee S, Rana R, et al. Perinatal intervention strategies providing food with micronutrients to pregnant and breastfeeding women in low-and middle-income countries: A scoping review. Maternal & Child Nutrition. 2024:e13681.

57. Ngesa O, Mwambi H. Prevalence and risk factors of anaemia among children aged between 6 months and 14 years in Kenya. PloS one. 2014;9(11):e113756.

58. Nambiema A, Robert A, Yaya I. Prevalence and risk factors of anemia in children aged from 6 to 59 months in Togo: analysis from Togo demographic and health survey data, 2013–2014. BMC public health. 2019;19:1–9.

59. Ntenda PA, Nkoka O, Bass P, Senghore T. Maternal anemia is a potential risk factor for anemia in children aged 6–59 months in Southern Africa: a multilevel analysis. BMC public health. 2018;18:1–13.

60. Moschovis PP, Wiens MO, Arlington L, Antsygina O, Hayden D, Dzik W, et al. Individual, maternal and household risk factors for anaemia among young children in sub-Saharan Africa: a cross-sectional study. BMJ open. 2018;8(5):e019654.

61. Wedderburn CJ, Weldon E, Bertran-Cobo C, Rehman AM, Stein DJ, Gibb DM, et al. Early neurodevelopment of HIV-exposed uninfected children in the era of antiretroviral therapy: a systematic review and meta-analysis. The Lancet Child & Adolescent Health. 2022.

62. Gollwitzer ES, Marsland BJ. Impact of early-life exposures on immune maturation and susceptibility to disease. Trends in immunology. 2015;36(11):684–96.

63. Camacho-Morales A, Caba M, García-Juárez M, Caba-Flores MD, Viveros-Contreras R, Martínez-Valenzuela C. Breastfeeding contributes to physiological immune programming in the newborn. Frontiers in pediatrics. 2021;9:744104.

64. Belderbos M, Houben M, Van Bleek G, Schuijff L, Van Uden N, Bloemen-Carlier E, et al. Breastfeeding modulates neonatal innate immune responses: a prospective birth cohort study. Pediatric allergy and immunology. 2012;23(1):65–74.

65. Richard SA, McCormick BJ, Murray-Kolb LE, Patil CL, Chandyo RK, Mahopo C, et al. Characteristics associated with the transition to partial breastfeeding prior to 6 months of age: Data from seven sites in a birth cohort study. Maternal & child nutrition. 2021;17(3):e13166.

66. Ughasoro MD, Emodi I, Okafor H, Ibe B. Prevalence and risk factors of anaemia in paediatric patients in South-East Nigeria. South African Journal of Child Health. 2015;9(1):14–7.

67. Cornet M, Le Hesran J-Y, Fievet N, Cot M, Personne P, Gounoue R, et al. Prevalence of and risk factors for anemia in young children in southern Cameroon. The American journal of tropical medicine and hygiene. 1998;58(5):606–11.

68. Parbey PA, Tarkang E, Manu E, Amu H, Ayanore MA, Aku FY, et al. Risk factors of anaemia among children under five years in the Hohoe municipality, Ghana: a case control study. anemia. 2019;2019(1):2139717.

69. Cappellini MD, Comin-Colet J, de Francisco A, Dignass A, Doehner W, Lam CS, et al. Iron deficiency across chronic inflammatory conditions: International expert opinion on definition, diagnosis, and management. American journal of hematology. 2017;92(10):1068–78.

70. Carter RC, Georgieff MK, Ennis KM, Dodge NC, Wainwright H, Meintjes EM, et al. Prenatal alcohol-related alterations in maternal, placental, neonatal, and infant iron homeostasis. The American journal of clinical nutrition. 2021;114(3):1107–22.

71. Carter RC, Jacobson SW, Molteno CD, Jacobson JL. Fetal alcohol exposure, iron-deficiency anemia, and infant growth. Pediatrics. 2007;120(3):559–67.

72. Ross LJ, Wilson M, Banks M, Rezannah F, Daglish M. Prevalence of malnutrition and nutritional risk factors in patients undergoing alcohol and drug treatment. Nutrition. 2012;28(7-8):738–43.

73. Appiah PK, Nkuah D, Bonchel DA. Knowledge of and adherence to Anaemia prevention strategies among pregnant women attending antenatal care facilities in Juaboso District in Western-north region, Ghana. Journal of pregnancy. 2020;2020(1):2139892.

74. Onyeneho NG, Subramanian S. Anemia in pregnancy: Factors influencing knowledge and attitudes among mothers in southeastern Nigeria. Journal of Public Health. 2016;24:335–49.

75. Camaschella C, Nai A, Silvestri L. Iron metabolism and iron disorders revisited in the hepcidin era. Haematologica. 2020;105(2):260.

76. Mwangi MN, Mzembe G, Moya E, Braat S, Harding R, Robberstad B, et al. Protocol for a multicentre, parallel-group, open-label randomised controlled trial comparing ferric carboxymaltose with the standard of care in anaemic Malawian pregnant women: the REVAMP trial. BMJ open. 2021;11(11):e053288.

