## Supplementary Information for "Iron Deficiency Anaemia in Mothers and Infants from a South African Birth Cohort: Prevalence and Profile in the Context of Inflammation"

**This PDF file includes:**


**Table S1.** Classification of Child Anaemia by Age Across Study Visits

**Table S2.** Inflammatory Biomarker Concentrations for Mothers and Infants with Iron Metrics Across Study Visits

**Table S3.** Maternal and Infant Sample Characteristics According to BRINDA-Adjusted Antenatal Maternal Iron Deficiency Status

**Table S4.** Maternal and Infant Sample Characteristics According to BRINDA-Adjusted Postnatal Maternal Iron Deficiency Status at Study Visit 1 (±3-6 Months Postpartum)

**Table S5.** Maternal and Infant Sample Characteristics According to BRINDA-Adjusted Child Iron Deficiency Status In Infants with Serum Ferritin at Study Visits 2 and 3

**Table S1. Classification of Child Anaemia by Age Across Study Visits**

| Child Age at Haemoglobin Measurement | Haemoglobin Concentration Threshold for Anaemia [a] | Number (%) of Haemoglobin Measures | | | Number (%) of Anaemic Observations | | |
|---|---|---|---|---|---|---|---|
| | | Study Visit 1 (*n* = 195) | Study Visit 2 (*n* = 173) | Study Visit 3 (*n* = 99) | Study Visit 1 (*n* = 100) | Study Visit 2 (*n* = 47) | Study Visit 3 (*n* = 20) |
| **0 – 3 days** | **<14g/dL** | | | | 0 | | |
| **3 days – 1 month** | **<15g/dL** | | | | 0 | | |
| **1 – 2 months** | **<11.5g/dL** | | | | 0 | | |
| **2 - 3 months** | **<9.4g/dL** | 31 (15.90) | | | 4 (4) | | |
| **3 months – 6 months** | **<11.1g/dL** | 164 (84.10) | 9 (5.20) | | 96 (96) | 5 (10.64) | |
| **6 months to 23 months** | **<10.5g/dL** | | 164 (94.80) | 99 (100) | 0 | 42 (89.36) | 20 (100) |
| **24 – 59 months** | **<11g/dl** | | | | 0 | 0 | |
| **60 months +** | **<11.5** | | | | 0 | 0 | |

[a] Reference ranges for children between 0 and 6 months were obtained from GSH/UCT Pathology Laboratory guidelines, Groote Schuur Hospital, National Health Laboratory Service (Western Cape); effective date 23 January 2003. WHO guidelines were used for children over 6 months.
SI conversion factor: To convert to haemoglobin grams per litre, multiply by 10.

**Table S2. Inflammatory Biomarker Concentrations for Mothers and Infants with Iron Metrics Across Study Visits**

| Inflammatory Biomarker | Measurement [a] | | | | |
|---|---|---|---|---|---|
| | Antenatal Maternal (*n* = 109) | Postnatal Maternal (*n* = 234) | Child Visit 1 (*n* = 196) | Child Visit 2 (*n* = 158) | Child Visit 3 (*n* = 96) |
| ***hs*CRP (mg/L)** [b] | 8.20 (10.28) [0.3 – 65.3] | 4.87 (7.02) [0.10 – 78.71] | 2.54 (6.92) [0.10 – 51.34] | 3.5 (9.41) [0.10 – 64.40] | 3.75 (9.42) [0.1 – 56.05] |
| **AGP (g/L)** [b] | 0.55 (0.23) [0.27 – 1.78] | 0.97 (0.34) [0.29 – 2.13] | 0.74 (0.32) [0.29 – 1.99] | 0.91 (0.34) [0.34 – 2.23] | 1.03 (0.55) [0.24 – 3.81] |

Abbreviations. *hs*CRP = highly sensitive C-Reactive Protein, AGP = Alpha-1Acid Glycoprotein; sTfR, soluble transferrin receptor
[a] Values for continuous variables are presented as: mean (standard deviation) [range].
[b] Missing Values: Antenatal maternal *hs*CRP (*n*=4), postnatal maternal *hs*CRP (*n* = 8), infant *hs*CRP (*n* = 6 at study visit 1, *n* = 5 at study visit 3), infant AGP (*n* = 3 at study visit 1, *n* = 2 at study visit 2, *n* = 3 at study visit 3).

**Table S3. Maternal and Infant Sample Characteristics According to BRINDA-Adjusted Antenatal Maternal Iron Deficiency Status**

| Variable[a] | Total sample ($n$ = 109) | | |
|---|---|---|---|
| | Maternal Iron Deficiency [b] ($n$ = 42) | No Maternal Iron Deficiency [b] ($n$ = 67) | $p$ |
| **Maternal Characteristics** | | | |
| Adjusted serum ferritin (µg/L) [b,e] | 11.09 (2.51) [5.47 – 14.81] | 34.86 (23.61) [15.03 – 139.28] | <0.001*** |
| Adjusted sTfR (mg/L) [c,e] | 5.56 (2.3) [2.30 – 12.39] | 4.39 (1.34) [1.73 – 8.81] | 0.004** |
| *hs*CRP (mg/L) | 6.63 (7.83) [0.30 – 41.30] | 9.21 (11.52) [0.40 – 65.30] | 0.211 |
| AGP (g/L) | 0.50 (0.17) [0.29 – 0.91] | 0.59 (0.26) [0.27 – 1.78] | 0.064 |
| Monthly household income (ZAR) [f,g] | | | |
| <1000 | 8 (19.04) | 14 (20.90) | 0.979 |
| 1000-5000 | 18 (42.86) | 29 (43.28) | |
| 5000-10000 | 6 (14.29) | 13 (19.40) | |
| >10000 | 1 (2.38) | 1 (1.49) | |
| Education [f] | | | |
| Primary | 0 (0) | 2 (2.99) | 0.517 |
| Some secondary | 18 (42.86) | 30 (44.78) | |
| Completed secondary | 21 (50) | 27 (40.30) | |
| Some tertiary | 1 (2.38) | 6 (8.96) | |
| Completed tertiary | 2 (4.76) | 2 (2.99) | |
| Employed | 15 (35.71) | 21 (31.34) | 0.637 |
| Age at enrolment (years) | 28.10 (5.53) [19.6 – 40.3] | 29.16 (5.88) [19.0 – 40.4] | 0.351 |
| Food insecurity | 22 (52.38) | 38 (56.72) | 0.658 |
| COVID-19 effect on food insecurity [g] | | | |
| No disruption | 15 (35.71) | 25 (37.31) | 0.526 |
| Some disruption | 7 (16.67) | 11 (16.42) | |
| Extreme disruption | 10 (23.81) | 9 (13.43) | |
| Smoking during pregnancy [f] | 0 | 1 (1.49) | 1 |
| Alcohol during pregnancy [f] | 0 | 3 (4.48) | 0.283 |
| Depression during pregnancy | 12 (28.57) | 11 (16.42) | 0.130 |
| HIV infection during pregnancy [g] | 19 (45.24) | 25 (37.31) | 0.387 |
| **Infant Characteristics at Birth** | | | |
| Sex (male) [g] | 23 (54.76) | 23 (34.33) | 0.027* |
| Gestational age at birth (weeks) [g] | 39.63 (1.12) [37 - 42] | 38.95 (1.85) [33 - 41] | 0.092 |
| HIV infection [g] | 0 (0) | 0 (0) | n/a |
| Birth weight (g) [d,g] | 3283.33 (416.24) [2400 - 4360] | 3160.89 (522.82) [1780 – 4120] | 0.240 |
| Birth length (cm) [d,g] | 50.47 (2.80) [44 - 57] | 50.02 (3.35) [41 – 57] | 0.503 |
| Birth head circumference (cm) [d,g] | 35.07 (1.36) [32 – 38] | 34.46 (1.69) [30.5 – 37.5] | 0.075 |

*Abbreviations.* ZAR, South African Rand; HIV, Human Immunodeficiency Virus; COVID-19, Coronavirus Disease 2019; g, grams; cm, centimetres; *hs*CRP = highly sensitive C-Reactive Protein, AGP = Alpha-1Acid Glycoprotein; sTfR, soluble transferrin receptor.

[a] Values for continuous variables are presented as: mean (standard deviation) [range]. Values for categorical variables are presented as: count (%).

[b] Serum ferritin concentrations were adjusted for inflammation using the BRINDA regression correction approach. Antenatal maternal iron deficiency classified as adjusted serum ferritin concentrations <15µg/L

[c] sTfR concentrations were adjusted for inflammation using the BRINDA regression correction approach.

[d] The birth anthropometric measurements were conducted by trained labour staff in the ward. Infant length and head circumference were measured in cm to the nearest completed 0.5cm and weight was measured in kgs.

[e] Levene's test was significant. *T*-test results were interpreted based on equal variance not assumed.

[f] Fisher's exact test result interpreted due to one or more cells having an expected count of less than 5.

[g] Missing values: household income ($n$ = 19), effect of COVID-19 on food insecurity ($n$ = 32), maternal HIV ($n$ = 2), infant sex ($n$ = 16), infant gestational age ($n$ = 39), infant HIV ($n$ = 75), infant birth weight ( $n$ = 17), infant birth length ($n$ = 18), infant birth head circumference ( $n$ = 18).

**p* is significant at <0.05, ** *p* is significant at <0.01, ****p* is significant at <0.001.

**Table S4. Maternal and Infant Sample Characteristics According to BRINDA-Adjusted Postnatal Maternal Iron Deficiency Status at Study Visit 1 (±3-6 Months Postpartum)**

| Variable[a] | Total sample (*n* = 234 ) | | |
|---|---|---|---|
| | **Maternal Iron Deficiency** [b] **(*n* = 59)** | **No Maternal Iron Deficiency** [b] **(*n* = 175)** | ***p*** |
| **Maternal Characteristics** | | | |
| Adjusted serum ferritin (µg/L) [b,e] | 9.55 (3.11) [3.14 – 14.98] | 35.89 (20.28) [15.12 – 115.53] | <0.001*** |
| Adjusted sTfR (mg/L) [c] | 4.58 (1.45) [2.23 – 12.01] | 3.89 (1.11) [1.78 – 7.92] | <0.001*** |
| *hs*CRP (mg/L) [g] | 4.37 (5.49) [0.10 – 30.56] | 5.03 (7.46) [0.10 – 58.71] | 0.539 |
| AGP (g/L) | 0.94 (0.33) [0.29 – 1.87] | 0.97 (0.34) [0.38 – 2.13] | 0.538 |
| Monthly household income (ZAR) [f,g] | | | 0.476 |
| <1000 | 10 (16.95) | 33 (18.86) | |
| 1000-5000 | 27 (45.76) | 76 (43.43) | |
| 5000-10000 | 17 (28.81) | 39 (22.29) | |
| >10000 | 1 (1.69) | 11 (6.29) | |
| Education [f] | | | 0.575 |
| Primary | 2 (3.39) | 4 (2.29) | |
| Some secondary | 30 (50.85) | 78 (44.57) | |
| Completed secondary | 18 (30.51) | 72 (41.14) | |
| Some tertiary | 5 (8.47) | 13 (7.43) | |
| Completed tertiary | 4 (6.78) | 8 (4.57) | |
| Employed [g] | 15 (25.42) | 65 (37.14) | 0.090 |
| Age at study visit 1 (years) [g] | 28.61 (5.85) [18 – 42] | 29.05 (5.50) [18 – 44] | 0.601 |
| Food insecurity | 37 (62.71) | 91 (52) | 0.153 |
| COVID-19 effect on food insecurity [g] | | | 0.957 |
| No disruption | 33 (55.93) | 97 (55.43) | |
| Some disruption | 12 (20.34) | 38 (21.71) | |
| Extreme disruption | 11 (18.64) | 36 (20.57) | |
| Smoking at enrolment [f] | 2 (3.39) | 7 (4) | 1 |
| Alcohol at enrolment [f] | 1 (1.69) | 12 (6.86) | 0.193 |
| Depression at enrolment | 9 (15.25) | 31 (17.71) | 0.664 |
| HIV infection | 23 (38.98) | 58 (33.14) | 0.415 |
| **Infant Characteristics at Study Visit 1** | | | |
| Age (months) [g] | 3.83 (0.81) [2.14 – 5.23] | 3.77 (0.78) [2.01 – 5.85] | 0.631 |
| Sex (male) | 21 (35.59) | 100 (57.14) | 0.004** |
| HIV infection [g] | 0 (0) | 0 (0) | n/a |
| Microcephaly [d,f,g] | 0 (0) | 1 (0.57) | 1 |
| Underweight [d,f,g] | 2 (3.39) | 5 (2.86) | 1 |
| Stunting [d,g] | 7 (11.86) | 23 (13.14) | 0.798 |

*Abbreviations.* ZAR, South African Rand; HIV, Human Immunodeficiency Virus; COVID-19, Coronavirus Disease 2019; g, grams; cm, centimetres; . *hs*CRP = highly sensitive C-Reactive Protein, AGP = Alpha-1Acid Glycoprotein; sTfR, soluble transferrin receptor.

[a]Values for continuous variables are presented as: mean (standard deviation) [range]. Values for categorical variables are presented as: count (%).

[b] Serum ferritin concentrations were adjusted for inflammation using the BRINDA regression correction approach. Iron deficiency classified as adjusted serum ferritin concentrations <15µg/L

[c] sTfR concentrations were adjusted for inflammation using the BRINDA regression correction approach.

[d] The anthropometric measurements were conducted by trained research staff. Child weight and length measurements were converted to z-scores based on age and sex using Anthro software for WAZ, HAZ, and HCZ. Infants were classified as underweight, stunted, or having microcephaly if they had z-scores of less than -2 SDs.

[e] Levene's test was significant. *T*-test results were interpreted based on equal variance not assumed.

[f] Fisher's exact test result interpreted due to one or more cells having an expected count of less than 5.

[g] Missing values: maternal *hs*CRP at study visit 1 ($n = 8$), maternal age at study visit 1 ($n = 2$), maternal employment ($n = 2$), household income ($n = 20$), effect of COVID-19 on food insecurity ($n = 7$), infant age at study visit 1 ($n = 52$), infant HIV ($n = 162$), infant microcephaly at study visit 1 ($n = 5$), infant underweight at study visit 1 ($n = 5$), infant stunting at study visit 1 ($n = 8$).

**p* is significant at <0.05, ** *p* is significant at <0.01, ****p* is significant at <0.001.

**Table S5. Maternal and Infant Sample Characteristics According to BRINDA-Adjusted Child Iron Deficiency Status In Infants with Serum Ferritin at Study Visits 2 and 3**

| Variable [a] | Study Visit 2 (±6-12 months; *n* = 158) | | | Study Visit 3 (±12-18 months; *n* = 96) [g] | | |
|---|---|---|---|---|---|---|
| | Child Iron Deficiency [b] (*n* = 10) | No Child Iron Deficiency [b] (*n* = 148) | *p* | Child Iron Deficiency [b] (*n* = 31) | No Child Iron Deficiency [b] (*n* = 64) | *p* |
| **Infant Characteristics** | | | | | | |
| Adjusted Serum Ferritin (μg/L) [b,e,g] | 9.44 (1.85)<br>[6.44 – 11.68] | 38.10 (25.64)<br>[12.10 – 200.71] | <0.001*** | 8.15 (2.66)<br>[2.55 – 11.88] | 29.38 (18.35)<br>[12.32 – 107.59] | <0.001*** |
| Adjusted sTfR (mg/L) [c,e,g] | 10.31 (3.29)<br>[7.10 – 17.43] | 6.80 (1.46)<br>[4.17 – 12.23] | 0.013* | 7.13 (3.54)<br>[1.84 – 21.12] | 5.69 (1.45)<br>[3.56 – 10.02] | 0.042* |
| *hs*CRP (mg/L) [g] | 2.19 (2.80)<br>[0.10 – 7.54] | 3.59 (9.70)<br>[0.10 – 64.4] | 0.651 | 3.20 (10.11)<br>[0.10 – 56.05] | 4.03 (9.12)<br>[0.10 – 52.83] | 0.692 |
| AGP (g/L) [e] | 0.99 (0.38)<br>[0.51 – 1.56] | 0.91 (0.34)<br>[0.34 – 2.23] | 0.513 | 0.96 (0.65)<br>[0.24 – 3.81] | 1.06 (0.50)<br>[0.34 – 2.85] | 0.435 |
| Age (months) [e,g] | 8.59 (1.30)<br>[6.15 – 10.55] | 8.53 (1.61)<br>[5.29 – 12.20] | 0.906 | 13.99 (1.02)<br>[12.43 – 15.88] | 14.23 (1.36)<br>[12.0 – 18.05] | 0.386 |
| Sex (male) [f,g] | 7 (70) | 73 (49.32) | 0.328 | 21 (67.74) | 35 (54.69) | 0.225 |
| HIV infection [f,g] | 0 (0) | 1 (67.57) | 1 | 1 (3.23) | 0 (0) | 0.341 |
| Microcephaly [d,f,g] | 0 (0) | 4 (2.70) | 1 | 0 (0) | 1 (1.56) | 1 |
| Underweight [d,f,g] | 0 (0) | 5 (3.38) | 1 | 1 (3.23) | 1 (1.56) | 0.548 |
| Stunting [d,f,g] | 0 (0) | 16 (10.81) | 0.60 | 2 (6.45) | 9 (14.06) | 0.495 |
| **Maternal Characteristics** | | | | | | |
| Monthly household income (ZAR) [f,g] | | | | | | |
| <1000 | 2 (20) | 24 (16.22) | 0.209 | 8 (25.81) | 13 (20.31) | 0.928 |
| 1000-5000 | 6 (60) | 70 (47.30) | | 15 (48.39) | 33 (51.56) | |
| 5000-10000 | 0 (0) | 36 (24.32) | | 6 (19.35) | 10 (15.63) | |
| >10000 | 1 (10) | 8 (5.41) | | 1 (3.23) | 3 (4.69) | |
| Education [f] | | | | | | |
| Primary | 0 (0) | 3 (2.03) | 0.806 | 1 (3.23) | 2 (3.13) | 0.276 |
| Some secondary | 6 (60) | 66 (44.59) | | 18 (58.06) | 27 (42.19) | |
| Completed secondary | 4 (40) | 57 (38.51) | | 8 (25.81) | 27 (42.19) | |
| Some tertiary | 0 (0) | 16 (10.81) | | 3 (9.68) | 4 (6.25) | |
| Completed tertiary | 0 (0) | 6 (4.10) | | 0 (0) | 4 (6.25) | |
| Employed [f,g] | 2 (20) | 48 (32.43) | 0.720 | 10 (32.26) | 24 (37.50) | 0.695 |
| Age at enrolment (years) [g] | 29.26 (5.64)<br>[19.7 – 36.2] | 29.23 (5.69)<br>[18.1 – 42.6] | 0.988 | 30.10 (5.83)<br>[18.9 – 38.5] | 28.67 (5.90)<br>[18.0 – 44.1] | 0.269 |
| Food insecurity [f,g] | 4 (40) | 84 (56.76) | 0.340 | 12 (38.71) | 41 (64.06) | 0.020* |
| COVID-19 effect on food insecurity [f,g] | | | | | | |
| No disruption | 3 (30) | 64 (43.24) | 0.217 | 10 (32.26) | 27 (42.19) | 0.022* |
| Some disruption | 1 (10) | 23 (15.54) | | 2 (6.45) | 13 (20.31) | |
| Extreme disruption | 4 (40) | 24 (16.22) | | 11 (35.48) | 9 (14.10) | |
| Smoking at enrolment [f,g] | 0 (0) | 6 (4.05) | 1 | 1 (3.23) | 6 (9.38) | 0.421 |

| | | | | | | |
|---|---|---|---|---|---|---|
| Alcohol at enrolment [f,g] | 1 (10) | 9 (6.08) | 0.490 | 2 (6.45) | 7 (10.94) | 0.713 |
| Depression at enrolment [f,g] | 3 (30) | 26 (17.57) | 0.393 | 6 (19.35) | 10 (15.63) | 0.649 |
| HIV infection [f,g] | 3 (30) | 57 (38.51) | 0.743 | 15 (48.39) | 28 (43.75) | 0.670 |

*Abbreviations.* ZAR, South African Rand; HIV, Human Immunodeficiency Virus; COVID-19, Coronavirus Disease 2019; g, grams; cm, centimetres; . *hs*CRP = highly sensitive C-Reactive Protein, AGP = Alpha-1Acid Glycoprotein; sTfR, soluble transferrin receptor.

[a]Values for continuous variables are presented as: mean (standard deviation) [range]. Values for categorical variables are presented as: count (%).

[b] Serum ferritin concentrations were adjusted for inflammation using the BRINDA regression correction approach. Iron deficiency classified as adjusted serum ferritin concentrations <12µg/L.

[c] sTfR concentrations were adjusted for inflammation using the BRINDA regression correction approach.

[d] The anthropometric measurements were conducted by trained research staff. Child weight and length measurements were converted to z-scores based on age and sex using Anthro software for WAZ, HAZ, and HCZ. Infants were classified as underweight, stunted, or having microcephaly if they had z-scores of less than -2 SDs..

[e]Levene's test was significant. T-test results were interpreted based on equal variance not assumed.

[f] Fisher's exact test result interpreted due to one or more cells having an expected count of less than 5.

[g] Missing values: BRINDA adjusted serum ferritin (*n* = 1 at study visit 3), infant *hs*CRP (*n* = 5 at study visit 3), infant AGP (*n* = 2 at study visit 2, *n* = 3 at study visit 3), infant adjusted sTfR (*n* = 3 at study visit 2, *n* = 4 at study visit 3), infant age (*n* = 7 at study visit 2, *n* = 3 at study visit 3), infant sex (*n* = 1 at study visit 3), infant HIV (*n* = 104 at study visit 2, *n* = 55 at study visit 3), infant microcephaly (*n* = 1 at study visit 3), infant underweight (*n* = 1 at study visit 3), infant stunting (*n* = 2 at study visit 2, *n* = 1 at study visit 3), household income (*n* = 11 at study visit 2, *n* = 7 at study visit 3), maternal employment (*n* = 1 at study visit 2, *n* = 2 and study visit 3), maternal age at enrolment (*n* = 1 at study visit 3), food insecurity (*n* = 1 at study visit 3), effect of COVID-19 on food insecurity (*n* = 39 at study visit 2, *n* = 24 at study visit 3), maternal PTE (*n* = 1 at study visit 3), maternal PAE (*n* = 1 at study visit 3), maternal depression at enrolment (*n* = 1 at study visit 3), maternal HIV (*n* = 1 at study visit 3).

*$p$ is significant at <0.05, ** $p$ is significant at <0.01, ***$p$ is significant at <0.001.
